# IgA-driven neutrophil activation underlies post-Zika severe dengue disease in humans

**DOI:** 10.1101/2025.02.11.25322002

**Authors:** Jaime A. Cardona-Ospina, Vicky Roy, Dorca E. Marcano-Jiménez, Sandra Bos, Elias Duarte, José V. Zambrana, Agamjot Bal, Boyeong K. An, Antonio Gregorio Dias, Julia Zhiteneva, Julia Huffaker, Carlos Montenegro, Guillermina Kuan, Marcos J. Ramos-Benitez, Angel Balmaseda, Galit Alter, Eva Harris

**Affiliations:** Division of Infectious Diseases and Vaccinology, School of Public Health, University of California, Berkeley, Berkeley, CA; Grupo Biomedicina, Facultad de Medicina, Institución Universitaria Visión de las Américas, Pereira, Colombia; Ragon Institute of MGH, MIT, and Harvard, Cambridge, MA; Department of Basic Sciences, Ponce Health Sciences University and Ponce Research Institute, Ponce, Puerto Rico; Sustainable Sciences Institute, Managua, Nicaragua; Department of Epidemiology, School of Public Health, University of Michigan, Ann Arbor, MI; Centro de Salud Sócrates Flores Vivas, Ministerio de Salud, Managua, Nicaragua; Laboratorio Nacional de Virología, Centro Nacional de Diagnóstico y Referencia, Ministerio de Salud, Managua, Nicaragua

## Abstract

The four dengue virus serotypes (DENV1-4) and the related Zika flavivirus (ZIKV) are major public health concerns worldwide. Primary immunity against ZIKV increases the risk of a subsequent severe DENV2 infection, presenting a significant challenge for developing safe and effective ZIKV vaccines. However, the mechanisms driving this phenomenon remain unclear. Leveraging our long-standing Pediatric Dengue Cohort Study in Nicaragua, we show that serum anti-NS1 IgA antibodies elicited after a primary ZIKV infection drive neutrophil activation and correlate with increased risk of subsequent severe DENV2 disease. Depletion experiments combined with e*x vivo* functional NETosis assays confirmed that anti-NS1 IgA antibodies drive neutrophil activation in dengue hemorrhagic fever/dengue shock syndrome (DHF/DSS). Moreover, increased neutrophil degranulation in paired serum samples obtained during the acute DENV2 infection from the same individuals correlated with IgA binding to DENV2 NS1 and preceded the development of vascular leakage. This finding was corroborated in an orthogonal hospital-based study. Thus, serum anti-NS1 IgA enhances neutrophil activation in severe dengue, with implications for prognostics, therapeutics, and vaccines.

## Main Text

The four serotypes of dengue virus (DENV1-4) and Zika virus (ZIKV) are closely related flaviviruses that co-circulate and constitute the most prevalent causes of arboviral disease globally^1^. DENV infections exhibit a clinical spectrum ranging from a self-limited febrile illness (dengue fever [DF]) to more severe, potentially fatal forms such as dengue hemorrhagic fever and dengue shock syndrome (DHF/DSS) characterized by thrombocytopenia, vascular leakage, and hemorrhagic manifestations^2^. Sequential infections with different DENV serotypes or ZIKV can either confer protection or increase the risk of subsequent severe disease^3–6^. This phenomenon, primarily driven by the humoral immune response, presents significant challenges for the development of safe and effective vaccines.

During sequential DENV infections, sub-neutralizing cross-reactive IgG antibodies targeting structural viral proteins can facilitate viral entry into myeloid cells expressing Fcγ receptors, increasing viral load through a phenomenon known as antibody-dependent enhancement (ADE)^7,8^. In addition, afucosylated IgG antibodies generated during the acute secondary infection further enhance disease severity by promoting cellular activation via FcγRIIIA^9,10^. Epidemiological studies have shown that a primary ZIKV infection increases the likelihood of subsequent symptomatic DENV infections and severe disease^4,5^. This association is strongly correlated with antibody levels, as a specific range of ZIKV-induced DENV cross-reactive antibodies are linked to an elevated risk of severe outcomes upon a subsequent DENV infection^4^. Therefore, understanding antibody features that underlie increased risk is essential for developing safe and effective vaccines.

To explore this, we employed an agnostic systems serology approach to characterize DENV cross-reactive antibodies after a prior ZIKV infection and before a subsequent symptomatic infection with DENV2, leveraging unique serum samples from our long-standing Pediatric Dengue Cohort Study (PDCS) in Nicaragua^11^. Our findings reveal that pre-existing IgA antibodies, particularly those targeting the nonstructural protein 1 (NS1), are key predictive features of DHF/DSS. Using systems serology and functional assays in paired pre-infection and acute-phase samples from the same individuals, we provide novel insights into the role of anti-NS1 IgA antibodies in mediating heightened neutrophil activation, a characteristic of children that progressed to severe disease. This insight reveals a previously unappreciated mechanism of IgA-mediated neutrophil activation in dengue disease that may underpin progression to severe disease in sequential flavivirus infections. These results have broad implications for understanding immune responses against flaviviruses in co-endemic regions and informing vaccine development. Moreover, they highlight the potential role of serum IgA antibodies in mediating neutrophil-mediated pathogenesis in infectious diseases.

## Results

### Pre-existing IgA antibodies differentiate severe and non-severe DENV infections

A prior ZIKV infection increases the risk of both symptomatic and severe disease during a subsequent DENV infection^4–6^. While this elevated risk has been linked to DENV cross-reactive antibodies induced by ZIKV^4^, the specific characteristics of these antibodies remain undefined. To address this gap, we employed an agnostic systems serology approach to evaluate the antigenic targets and isotypes of pre-existing antibodies associated with increased risk of DHF/DSS.

Our analysis focused initially on serum samples collected after primary ZIKV infection and before subsequent DENV2 infection from 24 children enrolled in the PDCS in Nicaragua. These children experienced a primary ZIKV infection in 2016, followed by a DENV2 infection in 2019–2020, which resulted in either DF (n = 14) or DHF/DSS (n = 10; Fig. 1A). The median time between plasma sample collection and the subsequent DENV2 infection was 7.16 months (interquartile rank (*IQR)*: 6.32–7.97) for individuals who developed DF and 7.84 months (*IQR*: 6.52–8.29) for those who developed DHF/DSS (p=0.7). Using a multiplexed bead-based assay^12^, we characterized the antibody binding profile against the envelope protein (E), domain III of E (EDIII), and NS1 of DENV1–4 and ZIKV, evaluating total IgG, IgG1–4, IgM, and IgA antibodies.

**Fig 1.**
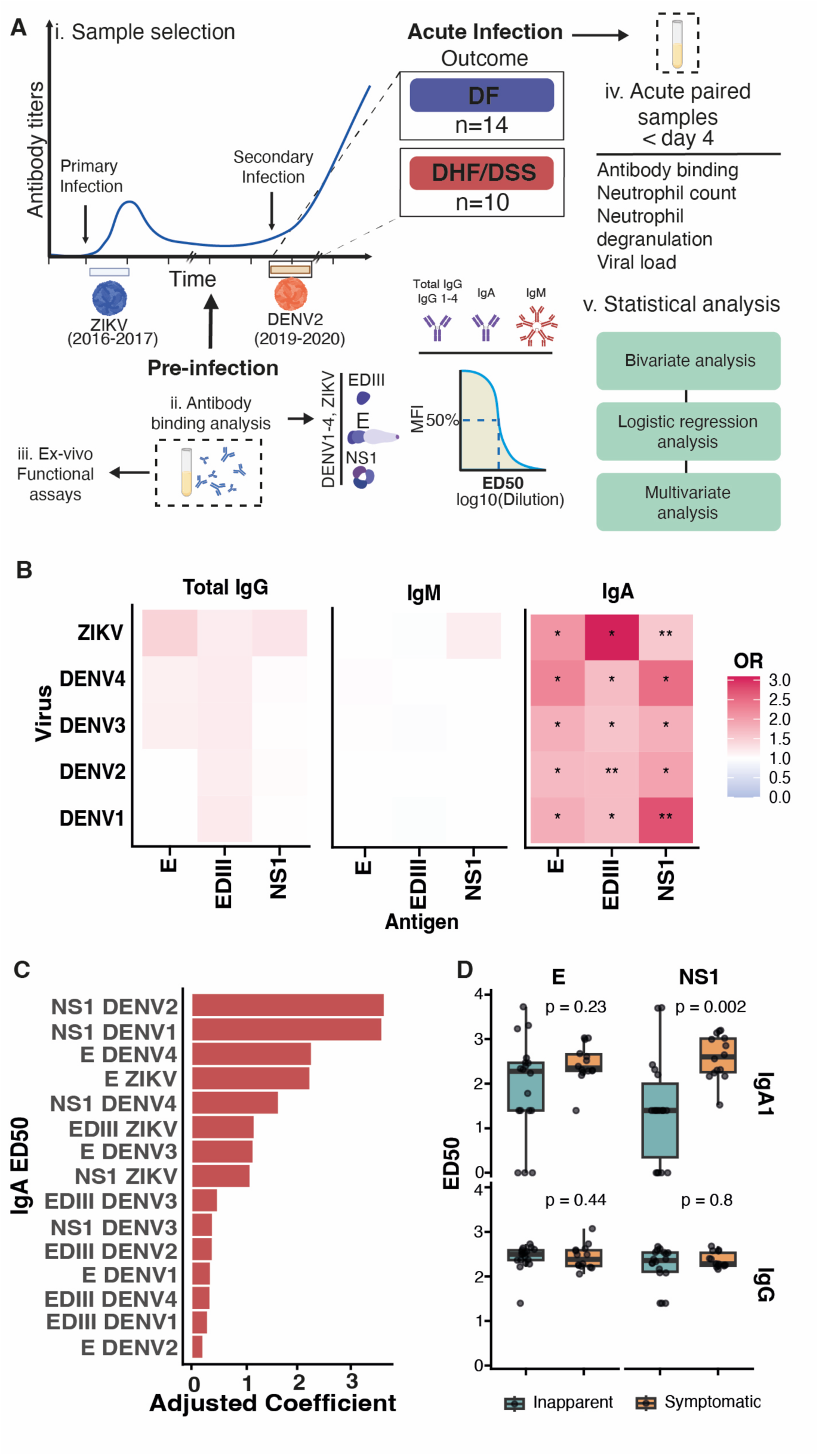
Study design and antibody profiling of post-ZIKV DENV2 cases in the Pediatric Dengue Cohort Study. (**A**) Study workflow. (i) Sample selection: plasma samples from children with primary ZIKV infection (2016) who developed dengue fever (DF, n=14) or dengue hemorrhagic fever/dengue shock syndrome (DHF/DSS, n=10) during a subsequent DENV2 infection (2019–2020) were analyzed. (ii) Antibody binding analysis was performed in pre-infection samples (after the primary ZIKV infection and before the subsequent DENV2 infection). (iii) *Ex vivo* functional assays with pre-infection antibodies were performed to analyze neutrophil activation, and (iv) neutrophil involvement during acute infection was assessed using paired acute-phase samples. (v) The statistical approach involved bivariate and multivariate analysis. (**B**) Heatmaps display the odds ratio of developing DHF/DSS for IgG, IgM, and IgA binding to the antigens tested. Significance: *p<0.05, **p<0.01 after False Discovery Rate (FDR) correction. (**C**) Adjusted coefficients via regularization reveal NS1 DENV2 binding as the strongest predictor of disease severity. (**D**) Comparison of ED_50_ values for IgA1 and IgG binding to E and NS1 antigens following primary ZIKV infection and prior to subsequent DENV2 infection, stratified by inapparent versus symptomatic outcomes. Boxplots show the median, interquartile range, and whiskers extending to 1.5× the interquartile range; individual dots represent participant values. Statistical comparisons were performed using one-tailed Wilcoxon rank-sum tests.

After correcting for false discovery rate (FDR), only binding of IgA antibodies differed significantly between the clinical groups across all tested antigens and isotypes (Extended Data Fig. 1A). Additionally, increased ED_50_ values of IgA antibodies, reflecting the dilution at which binding is reduced to 50%, were uniquely associated with an elevated risk of developing DHF/DSS (Fig. 1B). In a regularized multivariate model, IgA binding to NS1 of the incoming serotype (DENV2) emerged as the variable with the highest adjusted coefficient, highlighting its strong association with severe disease outcomes (Fig. 1C).

To further assess the specificity of the IgA immune response, we measured total IgA levels in plasma. Total IgA concentrations were comparable across groups and were within the physiological range (DF: median = 1.2 mg/mL, IQR: 0.6–1.3 mg/mL; DHF/DSS: median = 1.1 mg/mL, IQR: 0.9–1.2 mg/mL; p = 0.88), indicating that the observed differences in anti-DENV IgA were not due to overall systemic IgA levels but were instead antigen-specific. We next examined the clinical relevance of this response by comparing children with primary ZIKV infection who subsequently experienced either inapparent (n = 18) or symptomatic (n = 14) DENV2 infection. In pre-infection serum samples, only IgA binding to NS1-DENV2 differed significantly between the two groups, with higher levels in the symptomatic group (Fig. 1D), further supporting the role of pre-existing anti-NS1 IgA1 as a marker of disease risk. Next, to corroborate the discriminative potential of IgA binding suggested by the ED_50_ analysis, we assessed avidity by measuring antibody binding in the presence of urea as a chaotropic agent using our multiplexed bead-based assay and compared pre-infection samples of those who subsequently developed DF vs DHF/DSS^13^. Although individual antigen comparison did not reveal significant differences between clinical groups (Extended Data Fig. 2A), a multivariate supervised approach to assess the predictive value of IgA and IgG avidity for DHF/DSS showed that IgA avidity alone predicted DHF/DSS with an accuracy of 92% (95% CI: 62%–99%, p = 0.014; fig. S2B, while IgG avidity neither predicted outcome nor enhanced the model’s performance (Extended Data Fig. 2B).

These findings establish pre-existing high-avidity IgA antibodies induced after a primary ZIKV infection as key discriminative features associated with the development of DHF/DSS during a subsequent DENV2 infection.

### Pre-existing anti-NS1 IgA antibodies drive neutrophil effector functions *ex vivo* in children who developed DHF/DSS

Although IgA antibodies have been mainly studied in the context of mucosal immunity, they constitute the second most abundant immunoglobulin isotype in human serum, following IgG1^14,15^. Neutrophils are the primary and most abundant effectors of IgA antibodies^16^. Notably, IgA-mediated neutrophil activation is thought to contribute to the pathogenesis of IgA-mediated immune diseases through formation of IgA-immune complexes^17^. Several IgA-mediated immune diseases share clinical features with DHF/DSS, such as thrombocytopenia, arthralgia, petechiae and kidney involvement^18^. Considering this, we investigated whether pre-existing IgA antibodies could enhance neutrophil effector functions and be associated with the development of severe dengue disease.

To evaluate the functional capacity of pre-infection antibodies elicited by prior ZIKV infection to activate neutrophils upon engagement with antigens from the incoming DENV2 serotype (NS1 and E), we evaluated antibody-dependent neutrophil effector functions in *ex vivo* functional assays. Considering that phagocytosis and NETosis are key neutrophil effector functions^19^, we implemented *ex vivo* functional assays to measure antibody-dependent neutrophil phagocytosis (ADNP) and NETosis as a readout of antibody-dependent neutrophil activation. For ADNP, beads were coupled with either E or NS1 from the incoming serotype (DENV2) at comparable antigen densities (Extended Data Fig. 3A). The beads were opsonized with the pre-infection plasma of the participants, and flow cytometry was used to assess neutrophil phagocytosis of the beads^20^. Our results revealed that children who subsequently developed thrombocytopenia exhibited significantly higher ADNP against NS1 compared to those who did not (Extended Data Fig. 3B). In fact, ADNP against NS1 consistently exceeded ADNP against E across different clinical endpoints of disease severity (Extended Data Fig. 3B). This observation prompted a comparison of ADNP responses to NS1 vs. E among participants who developed DF or DHF/DSS and among participants who developed features of severe disease such as thrombocytopenia, plasma leakage, or hemorrhage. We found that the plasma from children who developed DHF/DSS or displayed hallmarks of severe disease like plasma leakage or thrombocytopenia consistently induced higher ADNP against NS1 than to E (Fig. 2A). Moreover, ADNP to NS1 was correlated with peak hematocrit during the subsequent DENV2 infection, and this marker of plasma leakage was strongly correlated with lower platelet count (Extended Data Fig. 3C). Further, IgA avidity to NS1, but not IgG avidity, was positively correlated with ADNP to NS1 (Extended Data Fig. 3C). Considering these observations and given that IgA antibodies against DENV2 NS1 showed the highest adjusted coefficient in our multivariate model of antibody binding and were significantly different across the spectrum of DENV infection (Fig 1B, D and Extended Data Fig. 1A), we sought to determine whether the antibody-dependent neutrophil effector functions induced by pre-existing antibodies were specifically driven by IgA and directed to NS1. To do this, we employed a complementary experimental approach. Due to the technical challenges of purifying IgA from small plasma volumes, we compared the ability of IgA-depleted and mock-depleted plasma to induce NETosis in an *ex-vivo* functional assay^21^. In this assay, wells coated with either E or NS1 were incubated with IgA-depleted or mock-depleted pre-infection plasma, and the induction of neutrophil extracellular traps (NETs) was quantified using a fluorescence-based assay. Importantly, in the absence of antibodies, E and NS1 induced minimal NET formation (Extended Data Fig. 4). Nevertheless, NS1 induced robust NETosis in the presence of mock-depleted serum (Fig 2B). Upon IgA depletion, responses to E were heterogeneous with no difference between DF and DHF/DSS, with some participants showing increased and others decreased neutrophil activation (Fig. 2B, Supplemental Videos 1 and 2). However, the response to NS1 was consistently reduced by IgA depletion in those who progressed to DHF/DSS (Fig. 2C, Supplemental Videos 3 and 4). All together, these results demonstrate that pre-existing antibodies, particularly IgA targeting NS1, play a critical role in driving neutrophil effector functions, as evidenced by ADNP and NETosis in *ex vivo* functional assays.

**Fig. 2.**
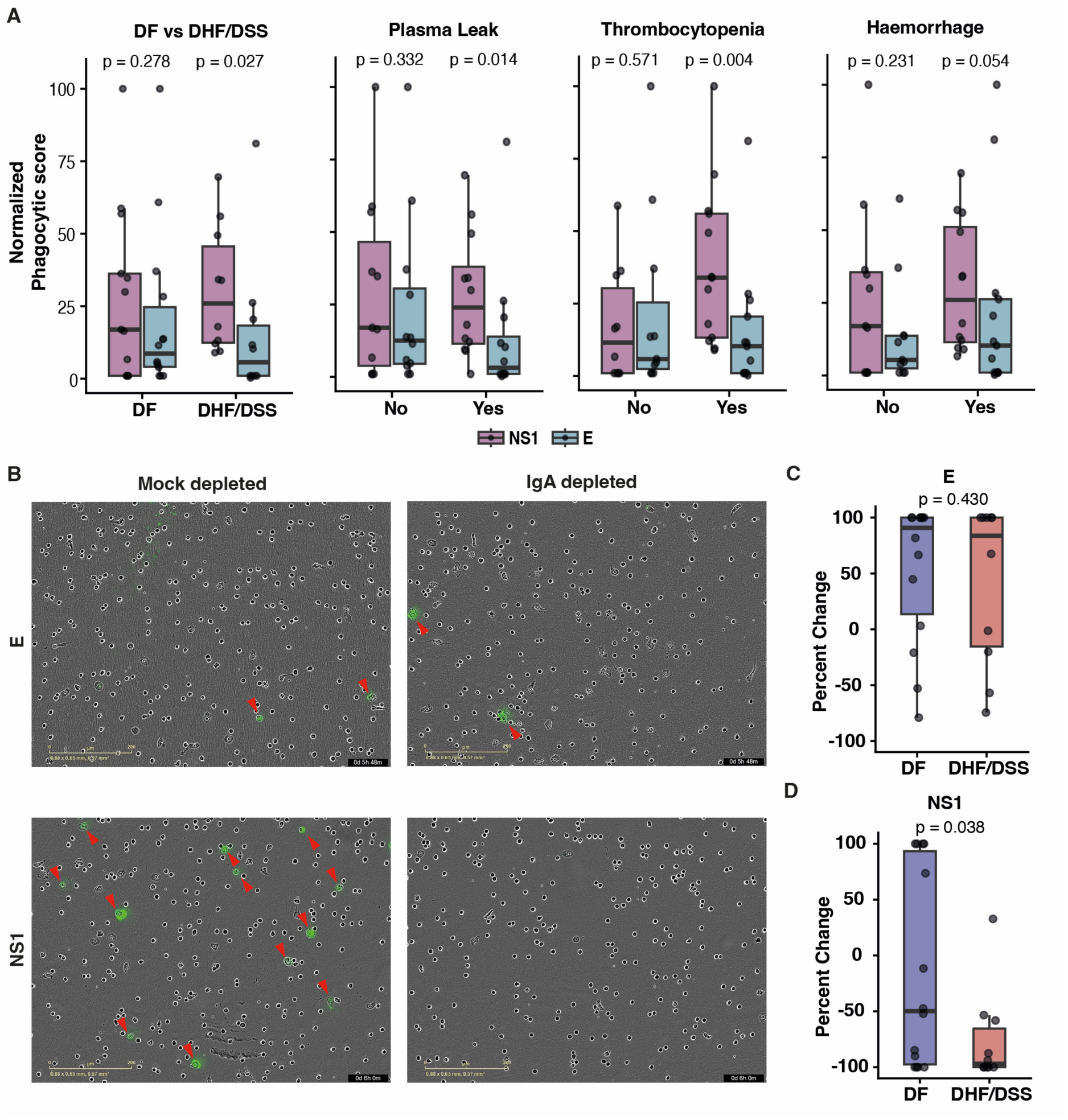
IgA antibodies targeting NS1 induce enhanced neutrophil activation in DHF/DSS cases. (**A**) Antibody-dependent neutrophil phagocytosis (ADNP) normalized phagocytic scores across clinical groups and severe disease manifestations. Data points represent the average of two replicates from two different healthy donors per sample (DF=14, DHF/DSS=10). Pre-infection plasma from children that subsequently developed DHF/DSS, thrombocytopenia, or plasma leakage exhibited significantly higher ADNP against NS1 compared to E, particularly in association with plasma leakage (p=0.014) and thrombocytopenia (p=0.004). The data indicate a skewed neutrophil activation response toward NS1 in severe cases. (**B**) Representative microscopy images of NETosis assays. Panels compare mock-depleted plasma (left) and IgA-depleted plasma (right) for E (upper) and NS1 (lower) antigens. IgA depletion significantly reduces NET formation in NS1-coated wells (p=0.038), highlighting the role of IgA in mediating neutrophil activation. Red arrows indicate NETs. Scale bar=200 μm. (**C**, **D**) Quantification of percent change in NETosis signal for E (**C**) and NS1 (**D**) antigens after IgA depletion. Data points represent the percent change comparing paired depleted and mock-depleted samples from participants who developed DF (n=14) or DHF/DSS (n=10). While no significant difference was observed for E, NETosis was significantly reduced in DHF/DSS cases when NS1-specific IgA was depleted. Statistical comparisons were performed using one-tailed Wilcoxon rank-sum tests.

### Post-ZIKV severe DENV infections are characterized by enhanced neutrophil activation in acute-phase plasma correlating with IgA binding to NS1

Consequently, we evaluated neutrophil involvement during the subsequent acute DENV2 infection in participants who progressed to DHF/DSS vs. DF. To do this, we assessed plasma samples obtained during the acute DENV2 infection of the same 24 participants (DF=14, DHF/DSS=10) from the PDCS. Initially, we evaluated circulating neutrophil levels during acute infection. On day three post-fever onset, just before the emergence of severe disease, children who progressed to DHF/DSS exhibited significantly higher relative neutrophil counts compared to those who developed DF (Fig. 3, A and B).

**Fig 3.**
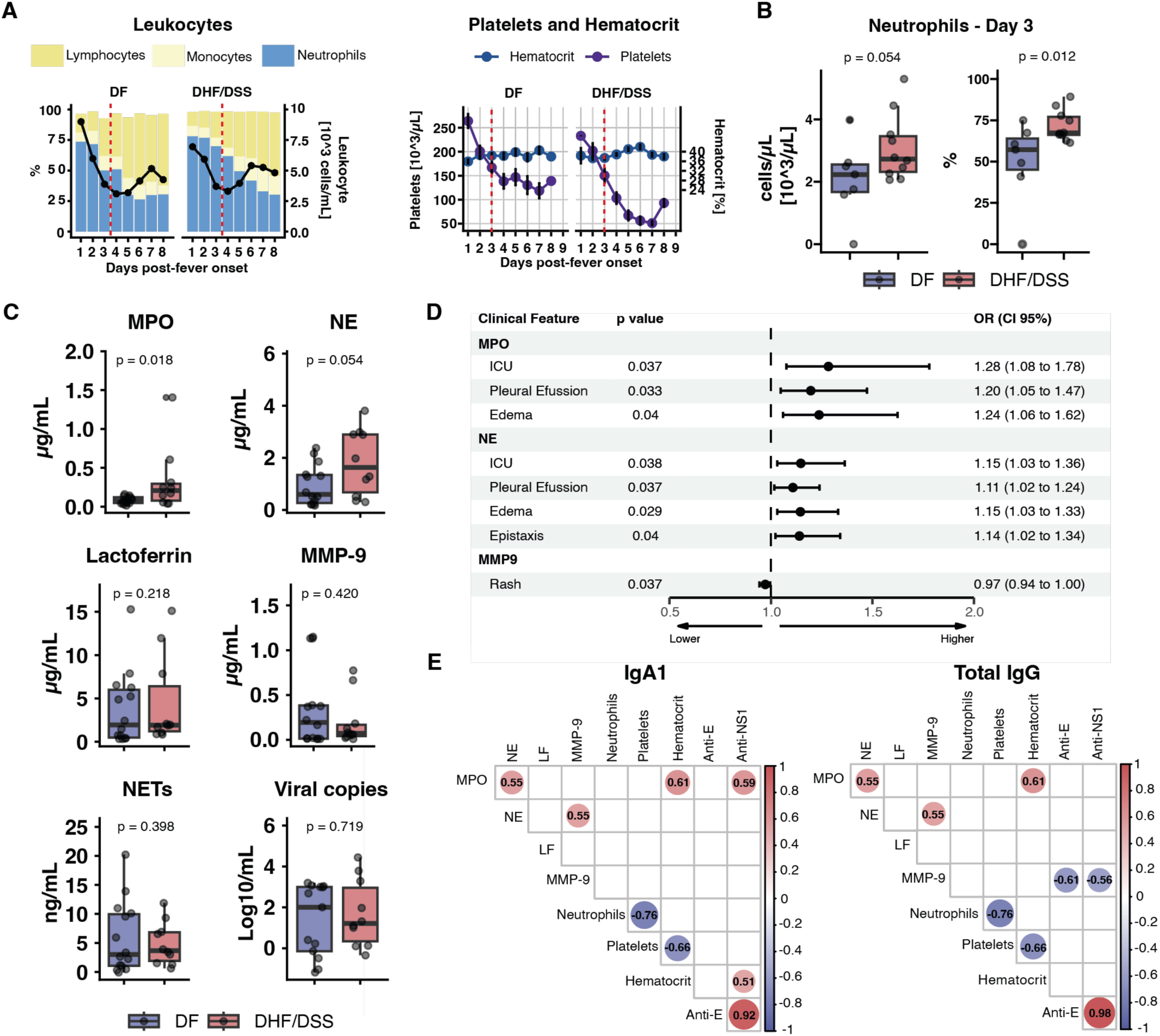
Enhanced neutrophil activation during acute DENV2 infection correlates with disease severity and binding of anti-NS1 IgA1 antibodies. (**A**) Relative leukocyte proportions (left panel), platelet counts, and hematocrit levels (right panel) in DF and DHF/DSS cases over time. Day 3 post-fever onset is marked with a dashed red line. The dark line corresponds to leukocyte count (units displayed on the right axis). (**B**) Neutrophil counts on day 3 post-fever onset—prior to the onset of thrombocytopenia (platelet count<100,000/µL)—were significantly elevated in children who progressed to DHF/DSS (p=0.012). Left panel: Absolute neutrophil count, Right panel: Relative neutrophil count. (**C**) Levels of primary granule markers (myeloperoxidase [MPO], neutrophil elastase [NE]), secondary granule markers (lactoferrin), tertiary granule markers (MMP-9), NETs, and viral loads among children with DF or DHF/DSS. DHF/DSS cases displayed higher levels of MPO (p=0.018) and NE (p=0.054), with no significant difference in viral load. In all boxplots, boxes represent the interquartile range (IQR) with median values shown as horizontal lines; whiskers extend to 1.5×IQR; individual points represent participants. Statistical comparisons were performed using one-tailed Wilcoxon rank-sum tests. (**D**) Logistic regression shows significant correlation between MPO, NE, and severe outcomes such as ICU admission and pleural effusion (OR with 95% CI). (**E**) Correlation heatmaps between antibody binding, neutrophil degranulation and clinical parameters of disease severity. Circle color indicates the direction of the correlation (red, positive; blue, negative), the circle size is proportional to the magnitude of the Pearson correlation coefficient, and the coefficient is displayed inside each circle. Only significant correlations (adjusted p-value<0.05) after FDR correction are displayed. IgA1 binding to NS1 positively correlates with MPO, while IgG inversely correlates with MMP-9. Anti-NS1 IgA1 positively correlates with hematocrit.

Next, we examined neutrophil degranulation, a key neutrophil effector function measurable in plasma. We analyzed levels of components of primary granules (myeloperoxidase [MPO], neutrophil elastase [NE]), secondary granules (lactoferrin [LF]), and tertiary granules (matrix metalloproteinase 9 [MMP-9]) in serum collected on days 1-3 post-fever onset (Table S1). Importantly, MPO levels were significantly elevated in children with DHF/DSS, and primary granule components (MPO and NE) were associated with heightened risk of developing severe clinical manifestations, including mucosal bleeding, vascular leakage, and increased risk of ICU admission (Fig. 3, C and D).

Thereafter, we evaluated whether this enhanced neutrophil degranulation was correlated with antibody binding. In agreement with our experimental observation and statistical prediction in pre-infection samples, IgA binding to NS1 positively correlated with levels of MPO, while IgG antibodies correlated negatively with levels of MMP-9 (Fig. 3E). Importantly, binding of anti-NS1 IgA positively correlated with higher hematocrit (Fig 3E). This highlights a distinct association between IgA antibodies targeting NS1, heightened neutrophil activation, and vascular leakage early in the evolution to severe disease.

In this study, analysis of DENV2 viral load during acute infection revealed no significant differences between clinical groups at the time of assessment (early post-fever onset; Fig. 3C), a finding also observed in post-ZIKV DENV infections in a different population in Brazil^6^.

These findings corroborate our experimental observations in pre-infection samples and *ex vivo* functional assays and demonstrate that severe post-ZIKV DENV2 infections are characterized by increased neutrophil activation that correlates with IgA antibodies in plasma binding to NS1. Furthermore, these results highlight plasma IgA antibodies directed against NS1 as key drivers of enhanced neutrophil activation during acute DENV2 infection.

### Enhanced neutrophil activation precedes the development of vascular leakage and progression to DHF/DSS

To validate our findings, we investigated neutrophil involvement in the Pediatric Dengue Hospital-based Study (PDHS), an independent study conducted in Managua, Nicaragua. This study enrolls participants under 15 years of age presenting with acute febrile illness suspected of dengue; DENV infections are laboratory-confirmed and serotyped by RT-PCR^22^. We screened children in the PDHS with DENV2 infections during the 2018–2020 epidemic to identify those with prior ZIKV exposure using our serotyping approach^4,12^.

A total of 111 dengue cases with past ZIKV infections were identified (DF=67, DHF/DSS=44). Consistent with the PDCS observations, children who developed DHF/DSS exhibited significantly higher neutrophil counts compared to those with DF prior to progression to DHF/DSS (Fig. 4A and Extended Data Fig. 5A). We further explored neutrophil activation in a subset of samples (18 DHF/DSS and 36 DF cases) collected early after fever onset (≤3 days post fever onset), matched for age and sex. As in prior analyses, significantly elevated markers of neutrophil degranulation, particularly MPO and NE components of primary granules, were observed in individuals who progressed to DHF/DSS (Fig. 4B). Importantly, neutrophil degranulation markers (MPO) were associated with anti-NS1 IgA binding, but not with IgA binding to E nor with IgG antibodies to NS1 or E (Fig. 4C and D). In addition, while anti-NS1 IgA levels positively correlated with hematocrit in the PDCS (Fig 3E), in the PDHS anti-E IgG antibodies were associated with lower hematocrit, a marker of lesser vascular leakage (Fig. 4C and D). Finally, we investigated whether neutrophil degranulation preceded the development of vascular leakage, a defining feature of DHF/DSS. To address this, we used gallbladder wall thickness as a quantitative marker of vascular leakage^23–25^. Gallbladder wall edema, a result of vascular leakage, is observed as DENV infection progresses to severe disease (Fig. 5A). This ultrasonographic metric provides a robust, continuous measure that is unaffected by treatment interventions and is widely and consistently employed in our setting for clinical triage and follow-up^23^. We found that the maximum gallbladder wall thickness – typically reached on day 4 post-fever onset or later (Fig. 5A) – was positively correlated with levels of the primary granule component NE measured on day 3 or earlier in both PDCS and PDHS participants (Fig. 5B and C). These findings reveal that neutrophil activation precedes vascular leakage, implicating a role in the pathogenesis of severe dengue disease.

**Fig. 4.**
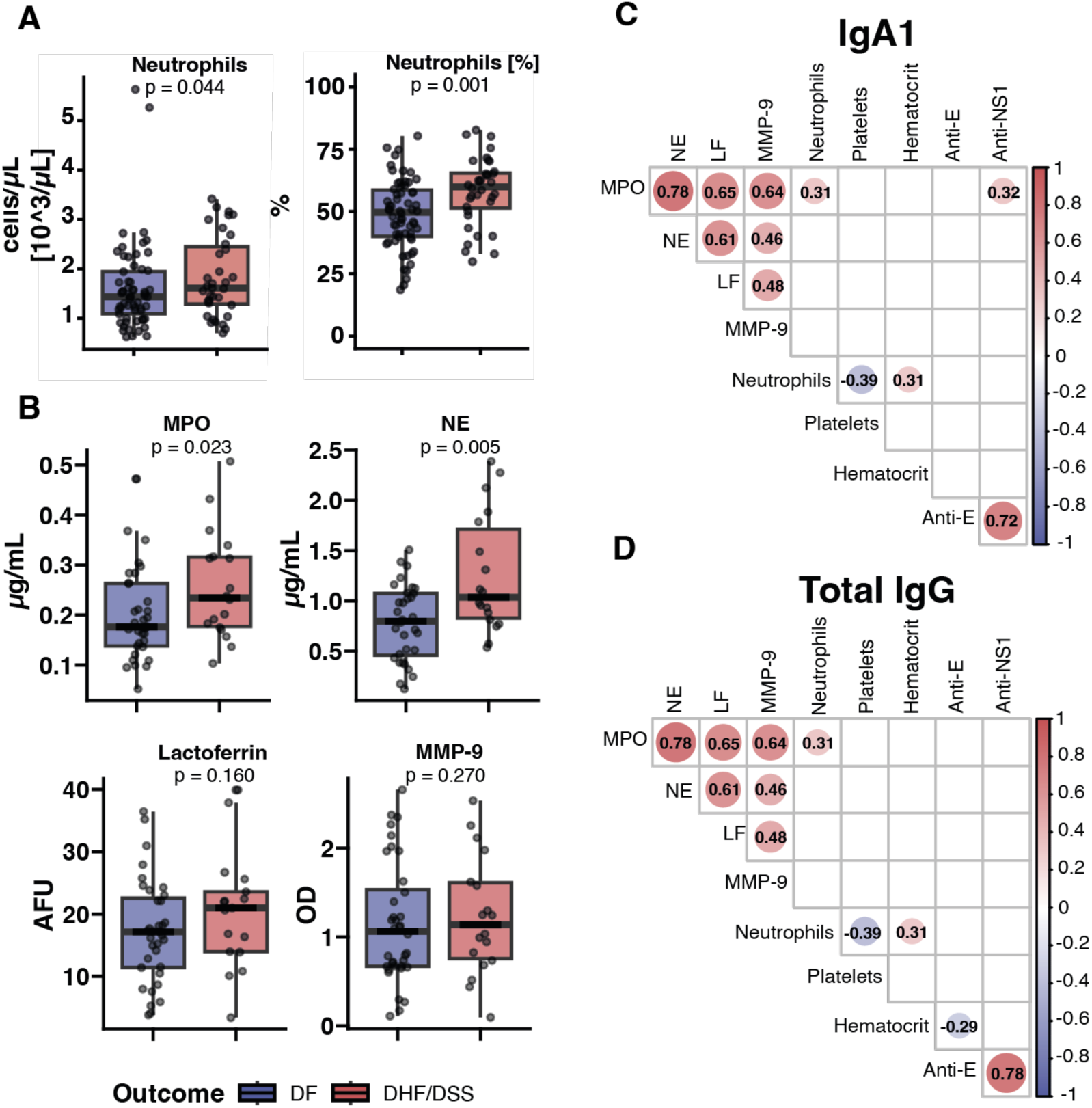
Neutrophil degranulation correlates with anti-NS1 IgA1 antibodies in an orthogonal hospital-based study. (**A**) Boxplots show the absolute count (left) and percentage (right) of circulating neutrophils in dengue fever (DF) and dengue hemorrhagic fever/dengue shock syndrome (DHF/DSS) cases. DHF/DSS was characterized by increased absolute (cells/uL) and relative (%) neutrophil counts. (**B**) Boxplots display plasma concentrations of neutrophil activation markers [myeloperoxidase (MPO), neutrophil elastase (NE), lactoferrin (LF), and matrix metalloproteinase 9 (MMP-9)] during acute DENV2 infection in each clinical group. In all boxplots, boxes represent the interquartile range (IQR) with median values shown as horizontal lines; whiskers extend to 1.5×IQR; individual points represent participants; p-values were calculated using one-tailed Wilcoxon’s rank-sum test. Participants who developed DHF/DSS had higher levels of primary granule components (MPO and NE). (**C, D**) Correlation of neutrophil relative counts, neutrophil degranulation and antibody binding with anti-NS1 and anti-E IgA1 antibodies (**C**) and Total IgG antibodies (**D**). Correlation heatmaps show pairwise Spearman’s correlation coefficients. Only significant correlations (p-value<0.05) are displayed. Circle color indicates correlation direction (red, positive; blue, negative), circle size is proportional to the absolute correlation coefficient, and the coefficient value is displayed within each circle. Anti-NS1 IgA1 antibodies correlate with increased MPO. Anti-E IgG negatively correlate with hematocrit.

**Fig. 5.**
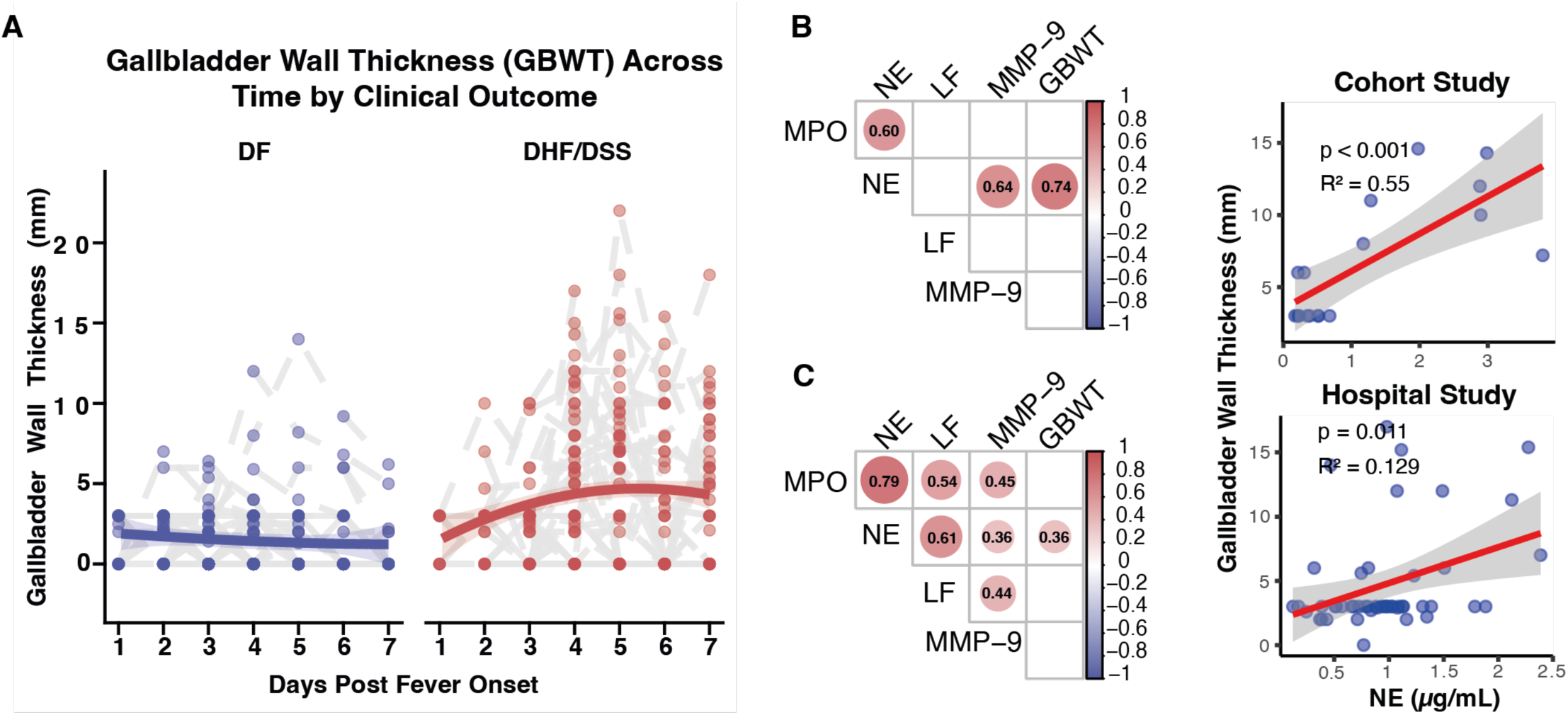
Neutrophil activation precedes and correlates with vascular leakage during acute DENV2 infection across two independent studies. (**A**) Gallbladder wall thickness (GBWT) over time in DF and DHF/DSS cases in the PDHS and PDCS. Greater gallbladder wall thickness was observed in DHF/DSS cases during acute infection, reflecting increased vascular leakage. (**B**, **C**) Correlation analyses between neutrophil degranulation and gallbladder thickness in the PDCS (**B**) and PDHS (**C**). Correlation heatmaps show pairwise Pearson’s correlation coefficients. Only significant correlations (p-value<0.05) are displayed. Circle color indicates correlation direction (red, positive; blue, negative), circle size is proportional to the absolute correlation coefficient, and the coefficient value is displayed within each circle. A positive correlation is observed between NE levels (measured on/before day 3 post-fever onset) and maximum gallbladder wall thickness in the PDCS (p<0.001, R²=0.55) and PDHS (p=0.01, R²=0.129).

## Discussion

The introduction of ZIKV into endemic regions of Latin America resulted in the widespread co-circulation of ZIKV and the closely related flaviviruses, DENV1-4. ZIKV continues to circulate alongside the four DENV serotypes at low levels in Asia and sporadically in Latin America^26^. Like sequential DENV infections with different serotypes, prior ZIKV immunity increases the risk of subsequent severe dengue disease ^4,5^. However, we and others found that post-ZIKV DENV infections that progress to severe disease do not exhibit the elevated viral load associated with antibody-enhanced DENV infections^6^. This suggests that alternative immune mechanisms may contribute to the heightened risk of severe disease in these cases. Therefore, we performed an agnostic serological analysis of potential biomarkers correlated with the risk of subsequent severe dengue disease. Surprisingly, plasma IgA antibodies were key discriminative features between clinical groups. Thus, we focused on plasma IgA, an often-overlooked antibody isotype in immune responses to infectious diseases. Our findings demonstrate that plasma IgA antibodies targeting NS1 drive neutrophil activation in severe post-ZIKV dengue disease. Importantly, we establish a temporal relationship between pre-existing anti-NS1 IgA antibodies and heightened neutrophil activation during the subsequent acute infection. In addition, we show that this increased neutrophil activation not only precedes but also correlates with the development of vascular leakage – a critical hallmark of severe dengue disease.

IgA antibodies in human serum are present at concentrations comparable to IgG3 and IgG2^14,15^. Although extensively studied in mucosal immunity, the role of IgA antibodies in human serum – whether protective or pathogenic – has been largely overlooked in infectious diseases. This gap partly reflects limitations in mouse models, which naturally lack CD89 (Fc alpha receptor), a critical receptor for IgA on human myeloid cells^27^. Of note, elevated IgA immune complexes have been implicated in autoimmune diseases through neutrophil activation; these diseases share clinical features with dengue, such as rash, petechiae, thrombocytopenia, and kidney involvement^17,18,28^. In the context of infectious diseases, serum IgA antibodies can mediate virus neutralization^29^, and DENV IgA antibodies targeting structural components can block ADE and neutralize the virion^30^. In contrast, our study demonstrates that IgA antibodies against NS1, a non-structural protein that is secreted during flavivirus infection, induce neutrophil activation in DHF/DSS preceding vascular leakage. While previous studies have reported DENV-associated IgA nephropathy and elevated titers of IgA in children with DHF/DSS^31,32^, the absence of a temporal relationship between IgA responses and disease progression has hindered understanding of its role in DENV pathogenesis. By analyzing samples collected after primary ZIKV infection and prior to subsequent DENV2 infection, we establish a temporal link between pre-existing ZIKV-induced IgA antibodies targeting DENV NS1 and heightened neutrophil activation, development of disease, and progression to DHF/DSS.

Importantly, heightened neutrophil activation has consistently been associated with severe dengue disease in humans^33–35^. Indeed, genes associated with increased neutrophil functions have been proposed as prognostic markers of severe dengue^36,37^. Nevertheless, the link between neutrophil activation and pre-existing antibodies, a major risk factor for DHF/DSS, has remained elusive. Interestingly, DENV induces the production of immature neutrophils^38^, which predominantly synthesize primary granules containing MPO and NE – degranulation markers associated with DHF/DSS. In parallel, serum IgA, to a greater extent than IgG, has been shown to enhance neutrophil effector functions during viral infections^39,40^. Based on these observations and our results, we hypothesize that IgA/NS1 immune complexes drive heightened activation of immature neutrophils during infection, contributing to vascular dysfunction and organ damage. The pathogenic role of neutrophil activation has been well-documented in other infectious diseases, such as malaria and leishmaniasis^41,42^. Although the current understanding of DHF/DSS immunopathogenesis is centered on the role of IgG antibodies targeting structural components of the virion in the context of ADE^9,10^, we believe that enhanced neutrophil activation due to cross-reactive anti-NS1 IgA antibodies may contribute to disease pathogenesis in sequential DENV infections.

Nonetheless, our study has several limitations. First, our work was not intended to define the entire pathogenic cascade leading to severe disease, but rather to illustrate a plausible immunological pathway linking pre-existing IgA immunity to downstream neutrophil responses specific to the incoming serotype. Existing murine DENV disease models lack expression of the Fc alpha receptor (CD89) and exhibit IgA immunobiology distinct from humans^27^. This limited our ability to validate IgA-mediated neutrophil activation as a direct cause of vascular leakage and define specific neutrophil effector functions leading to severe disease. Instead, we investigated the functional capacity of human IgA antibodies elicited by prior ZIKV infection to activate neutrophils upon binding to NS1 or E antigens from the incoming DENV serotype. We conducted *ex vivo* functional assays using primary cells from healthy human donors, which demonstrated a strong association between IgA antibodies targeting NS1, neutrophil activation, and severe disease. We showed that IgA depletion in the presence of NS1 but not E significantly reduced neutrophil effector functions, underscoring the specificity of this response. Then, we validated this experimental observation in two independent human studies during acute DENV2 infection.

However, our findings should be interpreted in light of the limitations of plasma biomarkers of neutrophil effector functions. We measured neutrophil degranulation and circulating NETs *in vivo* during the early phase of acute DENV infection. Nevertheless, circulating NET remnants may not reliably reflect the extent of NET formation in tissue sites where pathology occurs. Additionally, the timing of sample collection—restricted to the early febrile phase—may well not be optimal for capturing differences in NETosis. Unfortunately, matched samples from later timepoints in both the PDHS and PDCS, when NETosis may be more pronounced, are not available. Although we could not delineate the entire pathogenic cascade leading to severe disease, we demonstrate a plausible immunological mechanism linking pre-existing serum IgA immunity targeting NS1 of the incoming serotype to downstream neutrophil responses. This temporal connection represents a key strength of our study.

In addition, our findings suggest distinct roles for IgA antibodies targeting NS1 versus structural components such as E. The observed discrepancy between the capacity of anti-NS1 and anti-E IgA antibodies to trigger neutrophil activation raises important mechanistic questions that warrant further investigation. While both antigens were used under similar coupling densities, our findings suggest that the differences in neutrophil activation are not due to intrinsic properties of the antigens themselves. This is supported by the observation that neither NS1 nor E alone triggered NET formation. A more plausible explanation lies in the properties of the immune complexes formed with polyclonal sera. Variations in antibody abundance, avidity, stoichiometry, and/or epitope specificity between anti-NS1 and anti-E antibodies could influence the configuration and density of Fc receptor engagement on neutrophils. These factors, in turn, may modulate the strength and nature of downstream signaling required for neutrophil activation. While our current data are consistent with this hypothesis, a detailed mechanistic dissection of immune complex architecture, Fc receptor cross-linking, and signaling dynamics remains beyond the scope of this study and will require future molecular and biophysical analyses. Future studies are needed to elucidate these mechanisms and their broader consequences in the immunopathogenesis of viral infections.

Our study has several significant implications, as we explored the pathogenic role of plasma IgA antibodies, a frequently overlooked antibody isotype in immune responses. Our findings highlight the critical interplay between antigenic target and antibody isotype in disease and protection. While IgA antibodies targeting E can counteract ADE from IgG against the structural components of the virion^30^ and IgG anti-NS1 antibodies confer protection against the virulent properties of NS1^43,44^, our results show that anti-NS1 IgA antibodies are implicated in severe disease. Furthermore, while we do not exclude IgG-mediated ADE, our observations support the presence of an alternative pathogenic pathway mediated by IgA antibodies and neutrophil activation in DENV infection. This highlights an interesting therapeutic avenue, since neutrophil functions can be targeted by approved and emerging drugs^45^. Moreover, our findings underscore the importance of evaluating isotype- and antigen-specific antibody profiling of humoral immune responses induced by DENV and ZIKV vaccines. While further validation across diverse geographic and demographic settings is needed, our results suggest that binding and avidity of anti-NS1 IgA antibodies may serve as predictive markers of subsequent severe dengue disease. Similarly, the assessment of neutrophil degranulation early during DENV infection could provide a valuable triaging tool for identifying patients at risk of DHF/DSS.

Finally, our findings enrich the current understanding of the role of pre-existing immunity in the immunopathogenesis of severe dengue disease. We leveraged a key strength of the PDCS —namely, its unique ability to enable immune profiling in both pre-infection and acute-phase plasma samples in the same participants — and validated our findings in acute-phase samples from the PDHS as an orthogonal study. Using binding assays, *ex-vivo* functional assays, and validation of our observations in paired pre-infection and acute samples as well as an orthogonal study, we show that IgA antibodies drive neutrophil activation that correlates with increased risk of severe disease and precedes vascular leakage. We underscore the importance of antibody isotypes beyond IgG, such as IgA, and their antigenic targets (NS1 vs. E), with implications for establishing biomarkers of risk, refining therapeutic strategies, and informing vaccine development. Further research into the precise mechanisms of IgA-mediated neutrophil activation is essential to fully unravel the pathways driving severe disease in the context of flaviviral and other infectious diseases.

## Materials and Methods

### Ethics statement

The Pediatric Dengue Cohort Study was reviewed and approved by the institutional review boards of the University of California, Berkeley (protocol 2010-09-2245), the University of Michigan (HUM00091606), and the Nicaraguan Ministry of Health (NIC-MINSA/CNDR CIRE-09/03/07-008). The Pediatric Dengue Hospital-based Study was reviewed and approved by the institutional review boards of the University of California, Berkeley (2010-06-1649) and the Nicaraguan Ministry of Health (NIC-MINSA/CNDR CIRE-01/10/06-13). Parents or legal guardians of all subjects provide written informed consent, and subjects ≥6 years old provide assent.

### Study design

The primary objective of this study was to identify antibody biomarkers associated with the risk of developing severe dengue after a primary ZIKV infection during a subsequent DENV2 infection. Serum samples were obtained from participants enrolled in the Pediatric Dengue Cohort Study (PDCS), as described below. These participants had a documented primary ZIKV infection in 2016 and then experienced a DENV2 infection in the 2019-2020 dengue season. During the DENV2 infection, children developed either dengue fever (DF, n=14) or dengue hemorrhagic fever/dengue shock syndrome (DHF/DSS, n=10). We evaluated serum samples collected after the primary ZIKV infection and 4-10 months before the secondary DENV2 infection in order to identify antibody biomarkers of risk, using our multiplexed bead-based approach^12^, as described below.

The primary endpoint was the clinical classification of DF versus DHF/DSS, based on the WHO 1997 guidelines (see below). Secondary endpoints included the presence of plasma leakage, hemorrhagic manifestations, and thrombocytopenia. We also used gallbladder wall thickness, assessed by ultrasonography, as a surrogate of vascular leak^23^.

Following the identification of humoral biomarkers correlated with clinical outcomes (i.e., IgA antibodies targeting the viral NS1 protein), we investigated their role in disease pathogenesis. Given that neutrophils are the most abundant and primary effectors of IgA antibodies^16^, we performed *ex vivo* functional assays to evaluate antibody-dependent neutrophil effector functions (see below). Additionally, we examined the involvement of neutrophils and their correlation with antibody binding using paired acute-phase samples from the same participants of the PDCS during the acute secondary DENV2 infection. Finally, the findings were further validated with an independent dataset from the Pediatric Dengue Hospital Study (PDHS), evaluating participants who experienced a DENV2 infection during 2018 to 2020 with a history of primary ZIKV infection.

### Description of studies

The **Pediatric Dengue Cohort Study (PDCS)** is an ongoing prospective cohort study initiated in 2004 and based at the Sócrates Flores Vivas Health Center (HCSFV), the primary health care facility for District II of Managua, and part of the Municipal Health System of Managua. Located adjacent to Lake Xoxitlan, District 2 encompasses neighborhoods spanning a spectrum of low to middle socioeconomic status. The HCSFV serves ∼62,000 residents across 18 neighborhoods within District II^46^. The illiteracy rate in this area is approximately 7%, and over 90% of the households have access to running water and a sewage system. All residents have access to electricity. The PDCS tracks approximately 4,000 active participants yearly and has followed ∼9,800 children since its initiation. To preserve the original age distribution of the cohort, approximately 250 two-year-old children, along with children aged 3-11 years as needed, are enrolled annually. Adjustments to the age limit were implemented in 2018, 2019, and 2022, extending the eligibility to 15, 16, and 17 years, respectively. Participants benefit from free, round-the-clock medical care provided by study physicians at the HCSFV. In cases requiring hospitalization, study staff facilitate patient transfer to the study hospital, the National Pediatric Reference Hospital (Hospital Infantil Manuel de Jesús Rivera). Children presenting with fever and, since July 2016, rash are screened for signs and symptoms of dengue, Zika, and chikungunya. Symptomatic infections are captured through “enhanced” passive surveillance, complemented with periodic home visits. For each suspected case, both acute (0-6 days post-symptom onset) and convalescent samples (14-21 days) are collected to confirm infection. Approximately 97% of cases present to the HCSFV within the first three days of illness, and convalescent samples are collected from 94% of cases (Kuan et al 2009 AJE). Additionally, the study collects annual healthy blood samples from all participants in March-April (previously in July before 2011), which enables the detection of inapparent arboviral infections^11^.

The **Pediatric Dengue Hospital-Based Study (PDHS)** is based at the Hospital Infantil Manuel de Jesús Rivera (HIMJR), Managua, Nicaragua (2005-present)^22,47^. Children are invited to participate if they: 1) present to the hospital 1-7 days post-onset of symptoms and 2) meet case definitions for dengue, Zika, and/or chikungunya, which consists of either i) ≥1 signs or symptoms, including headache, arthralgia, myalgia, retro-orbital pain, positive tourniquet test, petechiae, or signs of bleeding; or ii) rash with ≥2 signs/symptoms including fever, conjunctivitis, arthralgia, myalgia, or peri-articular edema^22^. Blood samples are collected at presentation (acute), daily for days 1-3, and at convalescence (14-28 days). Infections are defined as for the cohort using RT-PCR, virus isolation, and DENV, ZIKV, and CHIKV serological assays (detailed below). Clinical information (>150 variables) and fluid intake/output is collected every 12 hours, and, after systematic monitoring by a clinical supervisor, is digitized by double-data entry, with quality control checks performed daily and weekly. A standardized questionnaire is used to record the subject’s demographic and epidemiologic characteristics. As part of a longitudinal arm of the study, consenting participants return to provide healthy blood samples 3, 6, 12, and 18 months post-illness.

### Case definitions

**Primary ZIKV infection in the PDCS** was defined as DENV- and ZIKV-naïve participants who presented with fever or rash after July 2016 and were clinically classified as either: A) meeting the 1997 and/or 2009 WHO criteria for dengue, or B) having an undifferentiated febrile illness. Laboratory confirmation of ZIKV infection was performed as described below.

**Primary ZIKV infection in the PDHS** was defined as participants meeting criteria A or B during 2018–2019 and showing evidence of a monotypic ZIKV antibody response early in infection (<4 days post-fever onset), as defined in ^4^ or ^12^ (detailed below).

**Second DENV2 infection** was defined as children who met the criteria for primary ZIKV infection and subsequently developed a DENV2 infection during 2019–2020 in the PDCS or 2018– 2020 in the PDHS. Children experiencing a second DENV2 infections were classified as DF or DHF/DSS based on the criteria described below.

### Laboratory confirmation of ZIKV and DENV infections in the PDCS

All cases meeting definitions A or B were evaluated for acute ZIKV or DENV infections, with laboratory confirmation performed by: 1) real-time RT-PCR (rRT-PCR) and/or viral isolation from acute serum (and/or urine for ZIKV); 2) seroconversion detected via ZIKV or DENV IgM capture (MAC)-ELISA; and/or 3) seroconversion or a ≥4-fold increase in DENV- or ZIKV-specific iELISA titers in paired acute and convalescent sera^11,48^. The infecting virus (DENV or ZIKV) was determined based on rRT-PCR results^49,50^. RT-PCR-negative cases were classified as 1) ZIKV if seroconversion or a ≥4-fold rise in ZIKV-specific titers was detected, or, for arbovirus cases in 2016–2017, if identified as ZIKV-positive by a computational classification algorithm^51^, or 2) Flavivirus cases if seroconversion or a ≥4-fold increase was observed in both DENV and ZIKV assays. Flavivirus cases and cases with confirmed RT-PCR or viral isolation evidence of co-infection with DENV and ZIKV were excluded from the analyses. All secondary DENV2 infections were confirmed by rRT-PCR^49^.

### Inclusion and exclusion criteria

All participants from the PDCS who had a documented primary ZIKV infection in 2016 followed by a second DENV2 infection during 2019-2020, with the second infection classified as DHF/DSS according to WHO 1997 criteria, were included (n=10). Additionally, 14 participants with a similar infection history who developed DF, matched by sex and age, were selected as a comparison group using convenience sampling, as well as 18 participants with the same infections history who developed a second inapparent DENV2 infection. The entire sample set (DF vs. DHF/DSS) was used for analysis of neutrophil counts during infection, excluding 7 participants with missing neutrophil counts on day 3 post-fever onset (Fig. 3B).

In the PDHS, a total of 362 confirmed DENV2 infections were documented between 2018 and 2020. Among these, 261 cases were classified as DF and 65 as DHF/DSS. All 65 DHF/DSS and a convenience sample of 103 DF cases were serotyped to identify second DENV2 infections after a primary ZIKV, using the methods described in ^4^ and ^12^ (detailed below). A total of 111 participants with past ZIKV infections were identified (DF=67, DHF/DSS=44). This entire sample set was used for analysis of neutrophil counts during infection, excluding 2 participants with missing neutrophil counts (Fig. 4A).

For our orthogonal confirmation of neutrophil involvement during acute DENV2 infection, we determined the sample size using Cohen’s *d* effect size and power analysis^52^. Based on our observations in the PDCS, the estimated effect size was *d* = −0.9128 (95% CI: −1.8139, −0.0117), corresponding to a large effect size. To achieve a statistical power of 80% (β = 0.20) at a significance level of 0.05 (*α* = 0.05) for a one-tailed two-sample *t*-test (assuming normal data distribution), a minimum of 16 participants per group (DF vs. DHF/DSS) was required. Accordingly, to analyze the temporal relationship between neutrophil functions and progression to severe disease, we selected a subsample of 18 DHF/DSS cases with serum samples collected on day 3 post-fever onset or earlier. These cases were matched by sex and age with 36 DF cases whose serum samples were also collected during acute infection (day 3 post-fever onset or earlier). This dataset was used to compare neutrophil degranulation among clinical groups and to evaluate the correlation of neutrophil degranulation with the maximum documented thickness of the gallbladder wall. Participant characteristics are summarized in **Table S1**. The plot in Fig. 5A included 7 DF and 10 DHF/DSS with gallbladder thickness information from the PDCS, and 66 participants with DF and 43 participants with DHF/DSS from the PDHS.

### Definition of severe dengue disease

Severe disease in our study was defined as a composite outcome of children meeting the WHO 1997 criteria for Dengue Hemorrhagic Fever or Dengue Shock Syndrome. **Dengue Hemorrhagic Fever** was defined as experiencing at least one of each of the following criteria: fever plus 1) hemorrhagic manifestations defined as the presence of at least one of the following manifestations: i) positive tourniquet test, ii) ecchymosis, petechia, hematoma, purpura, iii) bleeding at injection site, hypermenorrhea or vaginal bleeding, subconjunctival hemorrhage, hematuria, hemoptysis, bleeding gums or nose, iv) hematemesis or melena; 2) thrombocytopenia defined as platelet count ≤100,000 platelets per mm^3^; 3) evidence of plasma leakage as a result of increased vascular permeability defined as i) <u>≥</u>20% hemoconcentration, ii) age- and sex-specific elevated hematocrit, iii) pleural effusion, iv) ascites, v) hypoproteinemia or hypoalbuminemia, or vi) edema. **Dengue Shock Syndrome** (1997 WHO criteria) was defined as cases meeting criteria for DHF and at least one of each of the following criteria: 1) i) age-specific hypotension or ii) narrow pulse pressure <20 mmHg), and 2) i) cold, clammy skin and restlessness or ii) rapid and weak pulse. Symptomatic participants infected with DENV2 who did not meet the criteria for severe disease were categorized as having **Dengue Fever (DF)**. Additionally, secondary endpoints for the study included the presence of hemorrhagic manifestations, thrombocytopenia, or plasma leakage, as defined by the established criteria.

### Viral load quantification

Viral RNA was extracted from virus particles using the QIAmp kit (QIAGEN). The PCR standard curve used for the quantification of DENV genome copies was obtained with an amplicon containing a synthetic cDNA encompassing a fragment of the NS5 region (nucleotides 9937– 10113) from DENV2 strain DENV2 (8971.12a1SPD2; Genbank; MZ008454.1). Absolute quantitation of viral RNA was determined by real-time quantitative RT-PCR (RT-qPCR) using the following primer and probe sequences: DENV2 forward (ACAAGTCGAACAACCTGGTCCAT), reverse (GCCGCACCATTGGTCTTCTC) and probe (FAM-CCAGTGGAATCATGGGAAGAAGTCC CA-TAMRA. The Verso 1-step RT-qPCR kit (Thermo Fisher Scientific) was used following the manufacturer’s instructions. Standard curves were prepared using seven 10-fold dilutions of a DENV2 (8971.12a1SPD2; Genbank; MZ008454.1) synthetic fragment. Reaction mixtures were run in a QuantStudio 3 thermocycler (Applied Biosystems), and the quantity values were used for analyses^53,54^.

### EDIII serotyping in the PDHS

The EDIII serotyping of early acute serum samples (<4 days post-fever onset) from the PDHS was conducted as described in^12^. Briefly, recombinant EDIII proteins for DENV1-4 and ZIKV (courtesy of Dr. Lakshmanane Premkumar, University of North Carolina [UNC], Chapel Hill) were site-specifically biotinylated and conjugated to avidin-coated MagPlex Luminex beads. These antigen-coupled microspheres were combined and incubated in 384-well plates with appropriately diluted plasma (four 4-fold dilutions starting at 1:50). Each well contained 500 microspheres per antigen and was incubated for 90 minutes at 37°C, shaking at 1200 rpm. Following incubation, the plates were washed with phosphate-buffered saline (PBS) containing 0.1% bovine serum albumin (BSA) and 0.02% Tween 20 using an automatic magnetic washer (Tecan Hydrospeed). Antigen-specific IgG binding was detected with phycoerythrin (PE)-conjugated mouse anti-human total IgG (SouthernBiotech #9040-09). Median fluorescence intensity (MFI) was measured using an iQue3 (Intellicyt) instrument, set to acquire at least 50 beads per bead region. DENV-naïve human serum was included in triplicate on each plate as a negative control. MFI values for EDIII-DENV1, EDIII-DENV2, EDIII-DENV3, EDIII-DENV4, and EDIII-ZIKV were transformed as the relative proportion of the highest MFI observed at a serum dilution of 1:800. Then, a heatmap with hierarchical clustering using Wald’s method was generated, identifying five distinct clusters corresponding to DENV1-4 and ZIKV. The prior infecting virus (DENV1-4 or ZIKV) was assigned based on the antigen with the highest MFI value and the grouping within the cluster analysis. Finally, an empirical cutoff value was applied to distinguish monotypic EDIII ZIKV responses^12^. Specifically, for a sample to be classified as past primary ZIKV infection, the second highest MFI signal should be lower than 20% of the maximum ZIKV EDIII signal. Prior ZIKV infections were successfully detected in acute-phase sera from the PDHS using a similar EDIII-based method in^4^.

### Multiplex bead-based antibody binding and avidity assay

Recombinant E and NS1 proteins from DENV1-4 and ZIKV were commercially acquired from Native Antigen Co. (United Kingdom). Recombinant EDIII of DENV1-4 and ZIKV were site-specifically biotinylated and provided as a gift by Dr. Premkumar (UNC, Chapel Hill). Antibody subclass and isotype binding in pre-infection samples was determined using a multiplex bead-based assay, as previously described^12^. Briefly, E and NS1 from DENV1-4 and ZIKV were randomly biotinylated according to the manufacturer’s instructions (EZ-Link™ Sulfo-NHS-LC-LC-Biotin, No-Weigh™ Format; Thermo Fisher Scientific) and desalted using Zeba Spin Columns (Thermo Fisher Scientific). The randomly biotinylated E and NS1 and the site-specifically biotinylated EDIII from DENV1-4 and ZIKV were conjugated to avidin-coated MagPlex Luminex beads. Immune complexes were formed in 384-well plates by mixing the appropriately diluted plasma (six 4-fold dilutions) with antigen-coupled microspheres for 90 minutes at 37°C, shaking at 1200 rpm. After incubation, plates were washed using an automatic magnetic washer (Tecan Hydrospeed) with PBS containing 0.1% BSA and 0.02% Tween 20. For avidity testing, samples were washed for ten minutes with urea 7M or with PBS. Afterward, plates were washed again with PBS containing 0.1% BSA and 0.02% Tween 20. Finally, antigen-specific subclass/isotype binding was detected using PE-coupled mouse anti-human detection antibodies against IgA (SouthernBiotech #2050-09), IgM (SouthernBiotech #9020-09), total IgG (SouthernBiotech #9040-09), IgG1 (Southern-Biotech #9052-09), IgG2 (SouthernBiotech #9060-09), IgG3 (Southern-Biotech #9210-09), and IgG4 (SouthernBiotech #9200-09). Fluorescence was acquired using an iQue3 (Intellicyt) machine (Sartorius), as described above. Antigen-specific antibody subclass/isotype binding was extracted as median fluorescence intensity (MFI). Then, a netMFI was calculated by subtracting the background binding of a DENV-naïve sample from each test sample. Then, we fitted a dose-response curve using the netMFI across six 4-fold serial dilutions of plasma to estimate the effective dilution 50 (ED_50_) of the binding for each tested antigen, antibody subclass/isotype, and participant using the *drda* package v2.0.3. For calculating the Avidity Index (AI), the area under the curve (AUC) of the binding after washing with Urea or PBS was estimated, and then compared using the formula AI = AUC[Urea]/AUC[PBS]. Antigen-specific binding in acute phase samples was performed with E, NS1, and E dimer from DENV2. The recombinant stabilized envelope E dimer of DENV2 was purchased from the UNC Protein Expression and Purification Core^55^. Binding during the acute phase was analyzed as the AUC of the binding curve fitted using a dose-response model^54^.

### IgA depletion from serum

IgA antibodies were depleted from serum using the CaptureSelect™ IgA-XL Affinity Matrix resin (Thermo Scientific™) with a spin column method. Briefly, disposable Mobicol spin columns were prepared by loading either the CaptureSelect™ IgA affinity resin for depletion or empty agarose beads for mock depletion. The resins were equilibrated with PBS (pH 7.4) prior to sample application to ensure optimal binding conditions.

For depletion or mock-depletion, human serum diluted 1:50 was applied to the column containing the IgA-specific resin or empty resin. The column was incubated on a rotating wheel at room temperature for 30 minutes to allow antibody binding. After incubation, the column was centrifuged at 700 × g for 1 minute to collect the flow-through, which contained the IgA-depleted or mock depleted serum. Antibody binding, IgA depletion, and mock-depletion was confirmed in validation samples using the previously described antibody binding and isotype characterization multiplex bead-based protocol. Depleted and mock-depleted samples were stored at −80°C until further analysis.

### Antibody-dependent neutrophil phagocytosis and NETosis

Functional neutrophil *ex vivo* assays were conducted using neutrophils from three independent healthy human donors: two donors for antibody-dependent neutrophil phagocytosis (ADNP) and one donor for NETosis. Neutrophils were stimulated with immune complexes formed from pre-infection serum antibodies and the antigen of interest, as detailed below.

ADNP was performed as previously described^20^. Antigens (NS1-DENV2 or E-DENV2, Native Antigen Co.) were biotinylated and coupled to yellow-green neutravidin beads (Invitrogen). Immune complexes were formed by mixing coupled beads with plasma. Plasma was diluted 1:25 in assay buffer (PBS with 1% BSA and 0.05% Tween-20), based on prior titration experiments optimizing the dynamic range of the assay while minimizing background signal, and incubated with beads for immunocomplex formation for 2 hours at 37°C. White blood cells were isolated from fresh peripheral blood from healthy donors by performing ammonium-chloride-potassium lysis of red blood cells. The isolated white blood cells were then added to 96-well plates at a concentration of 2.5 × 10^5^ cells/ml, and neutrophils were identified using anti-CD66b PacBlue (BioLegend, 305112). Fluorescence was acquired using an iQue3 machine. A phagocytic score was calculated using the following formula: (percentage of bead-positive cells) x (geometric mean of MFI of bead-positive cells)/10,000. The results were summarized as the mean of the two independent experiments for each serum sample, and the value was normalized using the min-max approach. For this normalization, each value was rescaled using the formula: Normalized value = (x−min)/(max-min), where *x* is the individual phagoscore value, and *min* and *max* are the minimum and maximum values observed across all samples in the assay per antigen.

For NETosis, neutrophils were isolated from whole blood collected in EDTA tubes from one healthy donor under protocol #2303138796A001 at the Ponce Health Sciences University and Ponce Research Institute in Puerto Rico, using the EasySep Direct Human Neutrophil Isolation Kit (Stemcell Technologies, Catalog #19666). Cells were resuspended in Gibco Opti-MEM Serum Reduced Media (ThermoFisher, Catalog #31985070). To generate fixed immune complexes (ICs), NS1 or E protein (2 μg/mL) was plated in a 96-well plate and incubated overnight at 4°C with medium (negative control), Phorbol myristate acetate (PMA) as positive control, and mock-depleted or IgA-depleted samples. After protein coating, wells were washed three times with 1X PBS and then incubated with a 1:25 dilution of serum in 1X PBS (100 μL per well) overnight at 4°C. Wells were then washed three times with sterile 1X PBS containing 0.05% Tween 20 and then one time with 1X PBS before adding neutrophils. Neutrophils (30,000 cells/well) were added to each well in 100 μL of Gibco Opti-MEM containing 250 nM Cytotox Green (Essen BioScience, Catalog #4633). Plates were placed in an Incucyte SX5 instrument (Sartorius AG, Göttingen, Germany) for live-cell imaging. Phase-contrast and green fluorescence images were captured at 20X magnification, with an exposure time of 300 milliseconds for green fluorescence. Images were acquired every 30 minutes for 6 hours, with two images per well from distinct regions. The Incucyte software was trained to differentiate between normal cells and those undergoing DNA release using representative images from non-stimulated and stimulated cells. For analysis, the software applied surface-fit background correction with a fluorescence threshold of 2.0 Green Calibrated Units (GCU), edge sensitivity of −25, and a minimum mean intensity of 100. Phase-contrast analysis used artificial intelligence (AI) confluence detection. The software automatically analyzed the images and calculated the total integrated green intensity (GCU x µm^2^/ image) per well to quantify DNA release associated with NETosis. Paired IgA-depleted and mock-depleted samples were compared for each antigen to calculate the percentage of signal loss due to IgA depletion. To ensure comparability across samples, the percentage of signal loss was determined at the timepoint of the slope of the positive control (after one hour of stimuli) using the formula: Percentage change in signal = (IgA-depleted - IgA mock-depleted)/IgA mock-depleted * 100. The results were expressed as percentages, ranging from −100% to +100%.

### Quantification of markers of neutrophil degranulation

Neutrophil degranulation and NETosis were assessed in serum immediately after thawing, using commercially available kits following the manufacturers’ protocols. Myeloperoxidase (MPO) and Matrix Metalloproteinase-9 (MMP9) concentrations in plasma were quantified using DuoSet™ ELISA kits (R&D Systems, DY3174 and DY911, respectively). Briefly, 96-well plates were coated with specific capture antibodies diluted in PBS (pH 7.2–7.4) and incubated overnight at room temperature. Plates were blocked with 1% BSA in PBS for 1 hour at room temperature. Samples and standards, diluted in PBS with 1% BSA (with 2% normal goat serum for MMP9), were added (100 µL/well) and incubated for 2 hours at room temperature. Detection antibodies conjugated to biotin were applied, followed by a 20-minute incubation with streptavidin-horse radish peroxidase (HRP). Signal development was achieved using tetramethylbenzidine (TMB), and the reaction was stopped with 2N sulfuric acid. Optical density (OD) was measured at 450 nm with background correction at 570 nm. Standard curves were generated using serial dilutions of recombinant MPO and MMP9, and sample concentrations were interpolated using linear regression.

Human neutrophil elastase (NE) concentrations in plasma were quantified using the Hycult Biotech commercial kit (HK319). Plasma samples were diluted and added to microtiter wells pre-coated with a human elastase-specific capture antibody. Standards and samples (100 µL/well) were incubated for 1 hour at room temperature. After washing four times with wash buffer, 100 µL of biotinylated tracer antibody was added, and the plate was incubated for another hour at room temperature. Following additional washes, 100 µL of streptavidin-peroxidase conjugate was applied, and the plate was incubated for 1 hour. Signal development was performed by adding 100 µL of TMB substrate and incubating for 30 minutes in the dark. The reaction was terminated with 2% oxalic acid, and absorbance was measured at 450 nm using a microplate reader.

Human lactoferrin levels in plasma were measured using the CatchPoint® SimpleStep ELISA® kit (ab229392, Abcam), which employs fluorescence detection. Plasma samples were processed following the manufacturer’s recommendations. Standards were prepared by serial dilutions from a reconstituted lactoferrin stock solution. Standards and samples (50 µL each) were added to pre-coated wells, followed by 50 µL of an antibody cocktail containing capture and detector antibodies. Plates were incubated for 1 hour at room temperature on a plate shaker set to 400 rpm and subsequently washed three times with Wash Buffer. Detection was performed by adding 100 µL of CatchPoint HRP Development Solution, containing Stoplight Red Substrate and hydrogen peroxide, followed by a 10-minute incubation in the dark. Fluorescence intensity was measured at 530 nm (excitation), 570 nm (cutoff), and 590 nm (emission) using a fluorescence plate reader.

Neutrophil extracellular traps (NETs) in plasma samples were quantified using the GENLISA™ Human NETs ELISA Kit (KBH4548, Krishgen BioSystems). Plasma samples were diluted in the Standard Diluent provided. A standard curve was generated using serial dilutions of the NETs standard stock solution. Samples and standards (100 µL each) were added to wells of a microtiter plate pre-coated with anti-NETs antibodies, followed by a 60-minute incubation at 37°C. After aspiration, 100 µL of Biotinylated NETs Antibody Working Solution was added, and the plate was incubated for another 60 minutes at 37°C. Wells were then washed four times with 1X Wash Buffer to remove unbound components. Next, 100 µL of Streptavidin-HRP Working Solution was added to each well and incubated for 30 minutes at 37°C. Following additional washes, 100 µL of TMB substrate was added to develop the signal during a 10-minute incubation in the dark at 37°C. The reaction was terminated by adding 100 µL of Stop Solution. Absorbance was measured at 450 nm using a microplate reader within 15 minutes of adding the Stop Solution.

A standard curve was generated by plotting absorbance or fluorescence values against known analyte concentrations, and sample concentrations were interpolated and adjusted for dilution factors. Background correction was performed by subtracting the blank (zero standard) signal from all optical density (OD) or fluorescence values. Samples exceeding the upper limit of quantification were reanalyzed at higher serum dilutions when possible; otherwise, data was analyzed as background-corrected OD or fluorescence values. All assays were conducted in duplicate, and results were reported as the mean of the technical replicates. Outliers were identified and removed using the Interquartile Range (IQR) method. Boundaries for identifying outliers were defined as follows: Lower Bound: Q1 – (1.5 × IQR); Upper Bound: Q3 + (1.5 × IQR). Observations with values outside these thresholds were considered outliers and excluded from further analysis.

### Statistical analysis

All statistical analyses were performed using R (v4.2.3, R Foundation for Statistical Computing). Differences in the ED_50_ of binding antibodies in pre-infection samples between children who subsequently experienced DF or DHF/DSS were analyzed using a two-tailed Wilcoxon rank-sum test with false discovery rate (FDR) correction for multiple comparisons. Logistic regression models were employed to evaluate the association between pre-existing antibody profiles and the subsequent development of DHF/DSS. Each antibody characteristic was analyzed in a separate regression model, with FDR correction applied for multiple hypothesis testing.

To adjust the coefficients of the antibody characteristics correlated with DHF/DSS, we performed a multivariate regression with regularization. To estimate the regularization parameters, the dataset was divided into training and testing sets (50-50 split). Optimal regularization parameters (gamma and alpha) were determined via bootstrapping (n=1000), maximizing the area under the curve (AUC) on the training set. Then, the optimal parameters were applied to re-estimate the coefficients for selected antibody features using the entire dataset. Regularization was implemented using the glmnet R package (v4.1-860).

To compare the predictive accuracy of Total IgA versus Total IgG antibody avidity in pre-infection samples for distinguishing the children who subsequently developed DF or DHF/DSS, a similar regularization approach was used. In addition, the variables in the final model were assessed using an orthogonal partial least squares discriminant analysis (OPLS-DA) to estimate the variance in the response variable explained by the model (R²Y), the predictive performance for new data (Q²), and the overall classification accuracy. Two models were compared: avidity of IgA antibodies alone, and avidity of IgG antibodies alone.

Normalized phagocytic scores, percentage changes in signal in the NETosis assay, relative neutrophil counts, and levels of degranulation markers were compared across clinical groups using a one-tailed Wilcoxon rank-sum test. Logistic regression was employed to assess the association between degranulation marker levels and severe disease manifestations, estimating odds ratios (ORs) and 95% confidence intervals (CIs) using the oddsratio package (v2.0.1). Linear regression was used to evaluate the relationships between degranulation markers and antibody binding profiles, as well as between degranulation markers and maximum gallbladder wall thickness. All statistical analyses were conducted at a significance level of 5%.

## Data Availability

All data associated with this study are present in the main text or supplementary materials. The associated code is available for reference on Zenodo. Reference code and datasets for antibody profiling and coefficient adjustment using regularization (DOI: 10.5281/zenodo.14837883), reference code for assessing the classification performance of IgA, IgG, and IgG/IgA avidity (DOI: 10.5281/zenodo.14837889), and source data for figures (DOI: 10.5281/zenodo.14837915). After securing approval from the UC Berkeley Committee for the Protection of Human Subjects, individual data for figure reproduction can be shared with external researchers. For data access arrangements, please contact E.H. at. Standard data and material transfer agreements govern all data used in this study.

https://zenodo.org/records/14837883?token=eyJhbGciOiJIUzUxMiJ9.eyJpZCI6IjY4OTQ3MWQxLTY0M2MtNDMyZC1iZjgyLWI0MTk4NzRlMWRlMyIsImRhdGEiOnt9LCJyYW5kb20iOiI4YTQ3OGRjZThjYjM0Mzg0ZWUxMjA4ODI0YTJkYWI5NSJ9.4S52YOGN8E-LaZAYritkS-rarbAH-G2RHqEemMnKBiVjRON20lef2f1cz_jkcKnBz4lsPBrCBB-gNHSa-zI9tQ

https://zenodo.org/records/14837889?token=eyJhbGciOiJIUzUxMiJ9.eyJpZCI6IjJjNGM3Y2FlLTI3NmQtNGYxMC04YTczLWZjNzI5NWU5OWIyNiIsImRhdGEiOnt9LCJyYW5kb20iOiIzZWQ1OTQxZjNmZmY5ODgwNjg2ZmRkZWQzZmRiNTUyOCJ9.7NoQRzBq0oenoVar1wct01e6tV1G_YbxZjCKK_y7li6PboabaMq-qbr_lE7yDZPOKoCiEpEApCWnes9d23Nk4w

https://zenodo.org/records/14837915?token=eyJhbGciOiJIUzUxMiJ9.eyJpZCI6Ijk0ODVkZjk4LThkNzItNDE0Ni1iNDEzLTc1ODcwMzcxODBlZSIsImRhdGEiOnt9LCJyYW5kb20iOiJhNmQyNzVlYjkyN2JhNTk5YTE4MjE1NjE2MzZkNWUyMiJ9.ZOgLgpDEMc16UaN1Om3jDfzwXpUqD9TrVu6I9PoQkH0tSy1xpNC5B2M0HgIHIWsYQNXP6W-FSIh9jZMef8OfuQ

## Acknowledgments

We thank the study personnel at the Centro de Salud Sócrates Flores Vivas, the Nicaraguan National Virology Laboratory, the Hospital Infantil Manuel de Jesús Rivera, and the Sustainable Sciences Institute in Nicaragua. We are particularly grateful to the study participants and their families. We would also like to thank Olivia Cuaresma for her technical support, Myha Hill for support in code development, and Dr. Jeffrey R. Strich and Kiana Allen for their contributions to prior development of the NETosis assay.

## Author contributions

Conceptualization: JACO, EH

Methodology: JACO, EH, SB, JVZ, VR, BB, MJRB, GA, KA

Investigation: JACO, ED, AB, DEMJ, VR, BB, JZ, JH, AGD, KA

Visualization: JACO

Funding acquisition: EH, GA, MJRB

Project administration: EH Supervision: EH

Writing – original draft: JACO, EH

Writing – review & editing: JACO, EH, SB, JVZ, AGD, VR, AB, MJRB, DEMJ, GA, JZ, JH, KA

## Supplementary Materials

Figs. S1 to S5

Table S1

---

**Extended Data Figure 1. Antibody binding profile of pre-infection plasma of children after a primary ZIKV infection and before subsequent DENV2 disease. (A)** Antibody binding profiles for E, EDIII, and NS1 antigens from DENV1-4 and ZIKV in plasma prior to dengue fever (DF = 14) or dengue hemorrhagic fever/dengue shock syndrome (DHF/DSS = 10) cases. Binding levels (ED_50_) are shown for total IgG, IgG1-4 subclasses, IgM, and total IgA in a log_10_ scale. DHF/DSS cases consistently displayed significantly higher IgA binding levels across all DENV serotypes and antigens compared to DF cases. Statistical comparisons were performed using two-tailed Wilcoxon rank-sum tests and p values were adjusted by multiple comparisons. False discovery rate (FDR)-adjusted p-values are shown above the plots for significant associations.

**Extended Data Figure 2. Antibody avidity of IgA and IgG antibodies against E, EDIII, and NS1, and multivariate discrimination of clinical outcomes.** A) Boxplots show avidity indices of IgA (top) and IgG (bottom) binding to E, NS1, and EDIII antigens from DENV1–4 and ZIKV in dengue fever (DF) and dengue hemorrhagic fever/dengue shock syndrome (DHF/DSS) cases. Boxes represent the interquartile range (IQR) with medians indicated by horizontal lines; whiskers extend to 1.5×IQR; individual dots represent participants. Avidity was assessed by comparing antigen binding after exposure to 7 M urea (chaotropic agent) versus PBS. Statistical comparisons were performed using two-tailed Wilcoxon rank-sum tests and p values were adjusted by multiple comparisons. No significant associations were identified between groups. (**B**) Orthogonal partial least squares discriminant analysis (OPLS-DA) of avidity features distinguishes DF (blue) from DHF/DSS (red) cases. Left: Total IgA avidity predicted clinical outcome (Accuracy = 0.92, 95% CI: 0.62–1.0, p = 0.015; R²Y = 0.715; p = 0.004; Q² = 0.473; p = 0.002), whereas total IgG avidity did not (Accuracy = 0.5, 95% CI: 0.21–0.79, p = 0.811; R²Y = 0.291, p = 0.276; Q² = −0.14, p = 0.379).

**Extended Data Figure 3. Antibody-dependent neutrophil phagocytosis (ADNP) mediated by pre-infection antibodies.** (**A**) Coupling efficiency of biotinylated E and NS1 proteins to streptavidin-coated beads. Coupling density was quantified by probing with anti-biotin-PE and measuring median fluorescence intensity (MFI). Bars represent mean log10 (MFI) ± 95% CI from six independent replicates, showing comparable coupling efficiency of E and NS1. (**B**) Normalized ADNP scores for NS1 and E elicited by pre-infection plasma from DF (n = 14) and DHF/DSS (n = 10) cases. Pre-infection antibodies from children who later developed thrombocytopenia induced significantly higher ADNP against NS1. Data points represent the average of two replicates across two independent healthy donors per sample. (**C**) Spearman correlation heatmap showing associations between ADNP (NS1 or E) and clinical parameters (minimum platelet count, maximum hematocrit). Circle color indicates correlation direction (red, positive; blue, negative); size and intensity reflect magnitude. Only significant correlations are displayed (p<0.05). ADNP to NS1 was positively correlated with higher hematocrit during subsequent DENV2 infection. (**D**) Spearman’s correlation between ADNP to NS1 or E and the avidity index (AI) of IgA and IgG against the corresponding antigens. Only NS1-specific IgA avidity was significantly associated with ADNP to NS1. Adjusted p values after false discovery rate correction are shown.

**Extended Data Figure 4. Neutrophil activation and functional ex vivo responses to E and NS1 antigens in the presence of pre-infection antibodies**. Representative images of NETosis assays. Neutrophils were incubated with medium alone (negative control) or phorbol 12-myristate 13-acetate (PMA; positive control) in the presence of E or NS1 antigens. Green fluorescence indicates extracellular DNA release characteristic of NET formation. Scale bars: 200 μm.

**Extended Data Figure 5. Longitudinal dynamics of leukocyte populations, platelets and hematocrit in the Pediatric Dengue Hospital-based Study (PDHS) and correlations between degranulation markers and gallbladder wall thickness in the Pediatric Dengue Cohort Study (PDCS) and PDHS. (A)** Temporal dynamics of leukocyte populations (lymphocytes, monocytes, and neutrophils) in DF and DHF/DSS cases (left panel). The dashed red line marks the cutoff of normal leukocyte counts. Platelet counts and hematocrit trajectories during acute infection are shown (right panel) in the PDHS. **(B)** Pearson’s correlation analysis in the PDCS between gallbladder wall thickness and neutrophil degranulation markers, including myeloperoxidase (MPO), lactoferrin, and matrix metalloproteinase-9 (MMP-9). Raw p-values (non adjusted by multiple comparison) are displayed. MPO showed a significant positive correlation with gallbladder wall thickness (p = 0.030, R² = 0.276). **(C)** Correlation analysis in the PDHS between gallbladder wall thickness and neutrophil degranulation markers.

