## Supplemental Material for "IgA-driven neutrophil activation underlies post-Zika severe dengue disease in humans"

**This PDF file includes:**

Extended Data Figures. 1 to 5

Tables S1

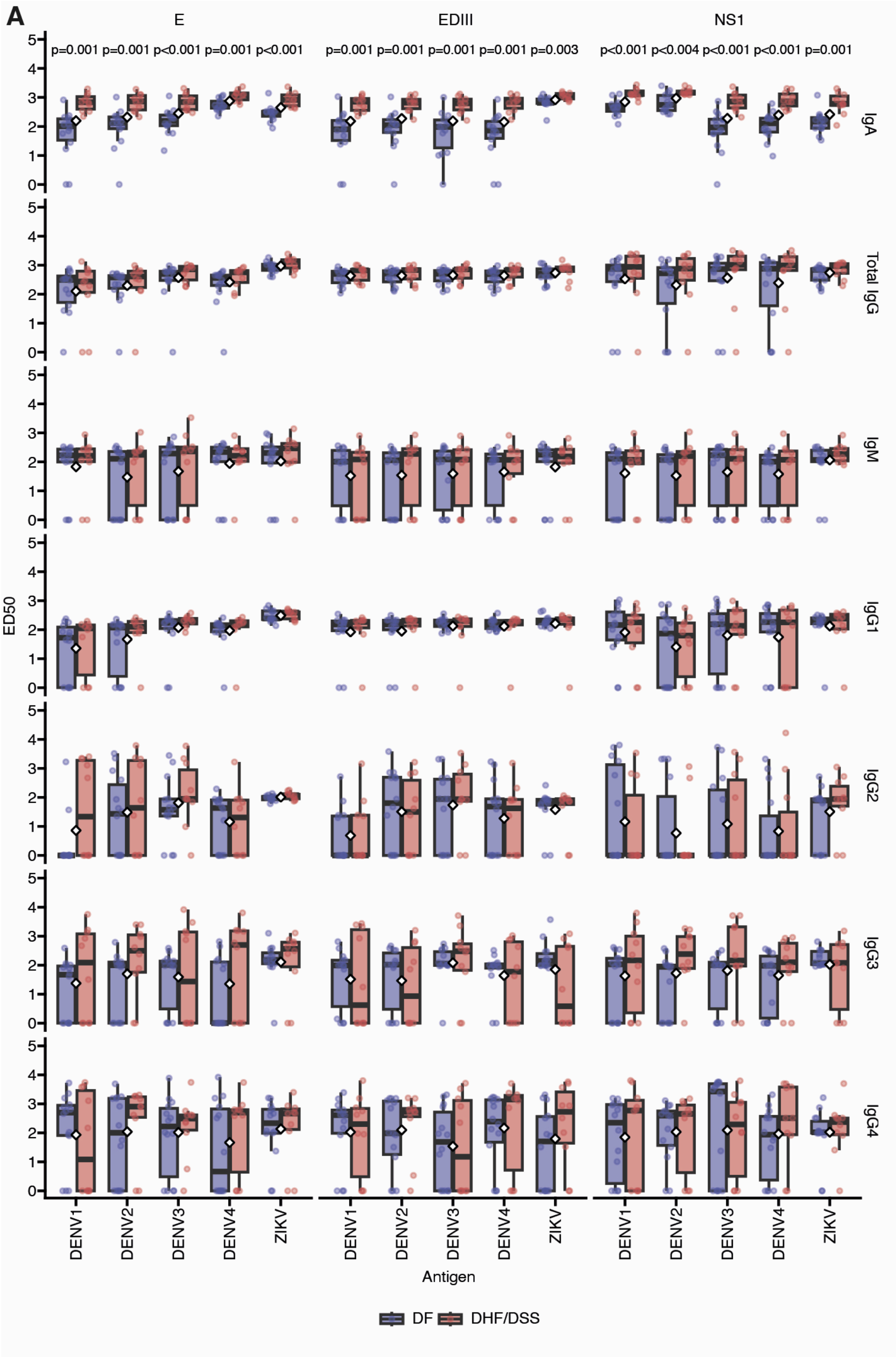


**Extended Data Figure 1. Antibody binding profile of pre-infection plasma of children after a primary ZIKV infection and before subsequent DENV2 disease. (A)** Antibody binding profiles for E, EDIII, and NS1 antigens from DENV1-4 and ZIKV in plasma prior to dengue fever (DF = 14) or dengue hemorrhagic fever/dengue shock syndrome (DHF/DSS = 10) cases. Binding levels (ED50) are shown for total IgG, IgG1-4 subclasses, IgM, and total IgA in a log10 scale.

DHF/DSS cases consistently displayed significantly higher IgA binding levels across all DENV serotypes and antigens compared to DF cases. Statistical comparisons were performed using two-tailed Wilcoxon rank-sum tests and p values were adjusted by multiple comparisons. False discovery rate (FDR)-adjusted p-values are shown above the plots for significant associations.

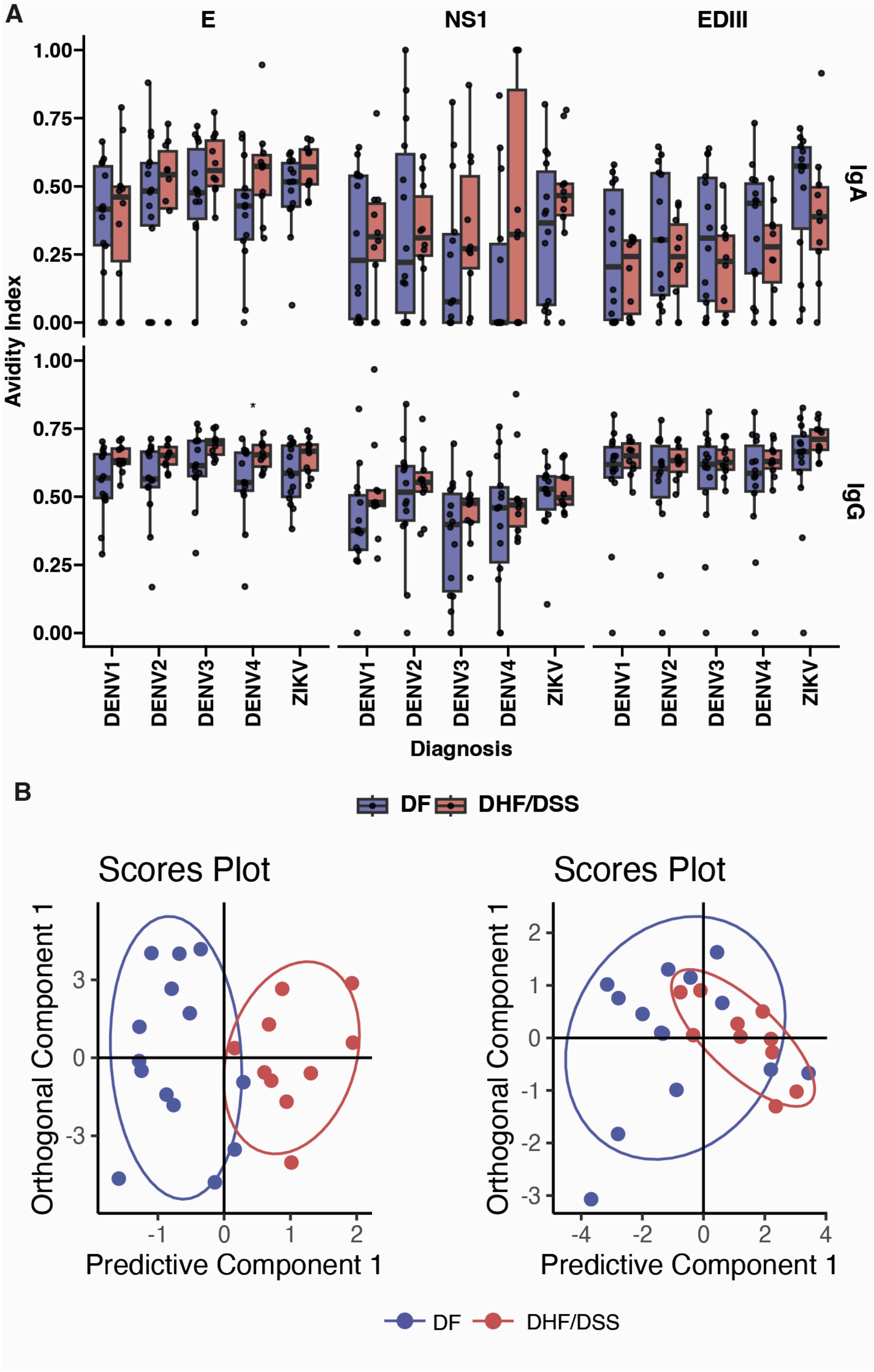


**Extended Data Figure 2. Antibody avidity of IgA and IgG antibodies against E, EDIII, and NS1, and multivariate discrimination of clinical outcomes.** A) Boxplots show avidity indices of IgA (top) and IgG (bottom) binding to E, NS1, and EDIII antigens from DENV1–4 and ZIKV in dengue fever (DF) and dengue hemorrhagic fever/dengue shock syndrome (DHF/DSS) cases. Boxes represent the interquartile range (IQR) with medians indicated by horizontal lines; whiskers extend to 1.5×IQR; individual dots represent participants. Avidity was assessed by comparing antigen binding after exposure to 7 M urea (chaotropic agent) versus PBS. Statistical comparisons were performed using two-tailed Wilcoxon rank-sum tests and p values were adjusted by multiple comparisons. No significant associations were identified between groups. (**B**) Orthogonal partial least squares discriminant analysis (OPLS-DA) of avidity features distinguishes DF (blue) from DHF/DSS (red) cases. Left: Total IgA avidity predicted clinical outcome (Accuracy = 0.92, 95% CI: 0.62–1.0, $p = 0.015$; $R^2Y = 0.715$; $p = 0.004$; $Q^2 = 0.473$; $p = 0.002$), whereas total IgG avidity did not (Accuracy = 0.5, 95% CI: 0.21–0.79, $p = 0.811$; $R^2Y = 0.291$, $p = 0.276$; $Q^2 = -0.14$, $p = 0.379$).

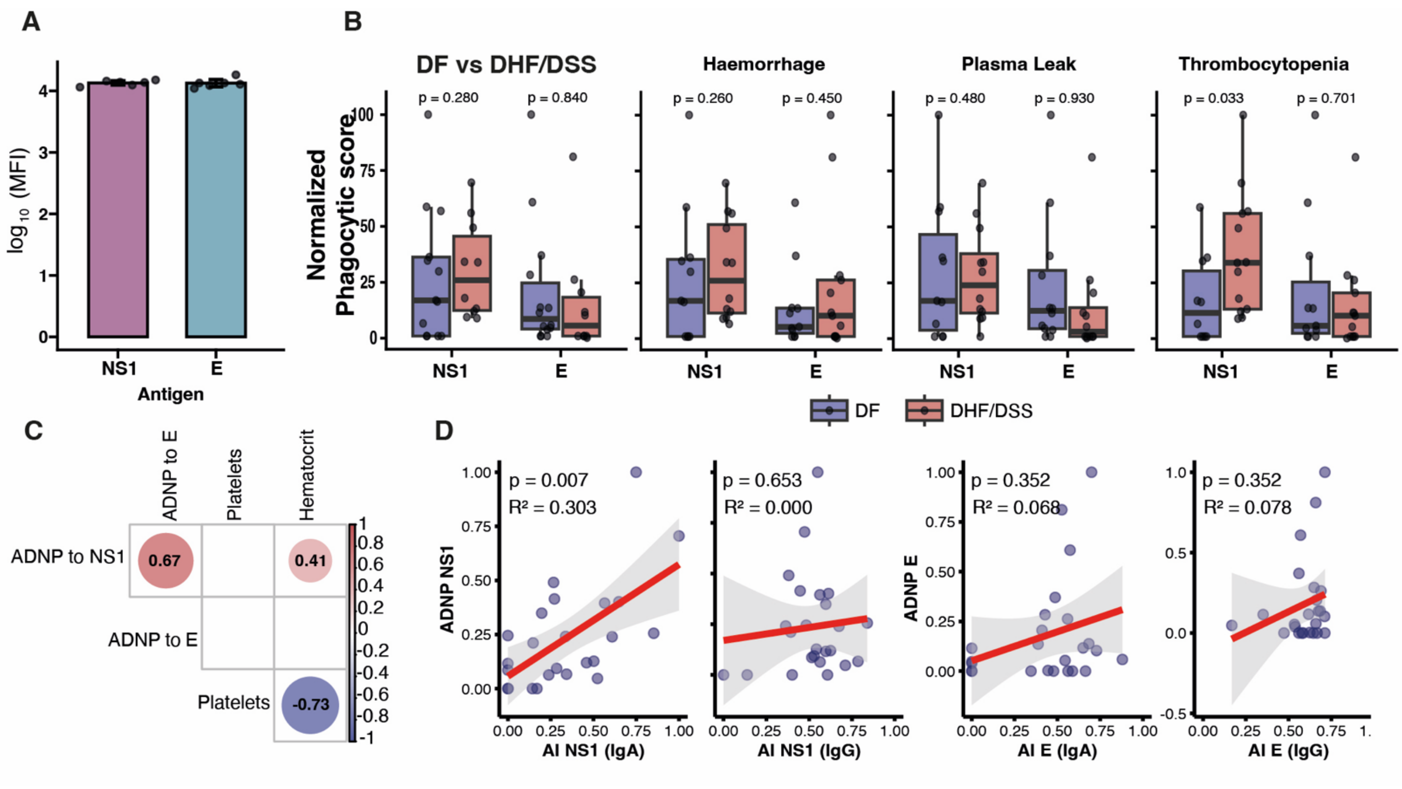


**Extended Data Figure 3. Antibody-dependent neutrophil phagocytosis (ADNP) mediated by pre-infection antibodies.** (**A**) Coupling efficiency of biotinylated E and NS1 proteins to streptavidin-coated beads. Coupling density was quantified by probing with anti-biotin-PE and measuring median fluorescence intensity (MFI). Bars represent mean $\log_{10}$(MFI) ± 95% CI from six independent replicates, showing comparable coupling efficiency of E and NS1. (**B**) Normalized ADNP scores for NS1 and E elicited by pre-infection plasma from DF ($n = 14$) and DHF/DSS ($n = 10$) cases. Pre-infection antibodies from children who later developed thrombocytopenia induced significantly higher ADNP against NS1. Data points represent the average of two replicates across two independent healthy donors per sample. (**C**) Spearman correlation heatmap showing associations between ADNP (NS1 or E) and clinical parameters (minimum platelet count, maximum hematocrit). Circle color indicates correlation direction (red, positive; blue, negative); size and intensity reflect magnitude. Only significant correlations are displayed ($p<0.05$). ADNP to NS1 was positively correlated with higher hematocrit during subsequent DENV2 infection. (**D**) Spearman's correlation between ADNP to NS1 or E and the avidity index (AI) of IgA and IgG against the corresponding antigens. Only NS1-specific IgA avidity was significantly associated with ADNP to NS1. Adjusted p values after false discovery rate correction are shown.

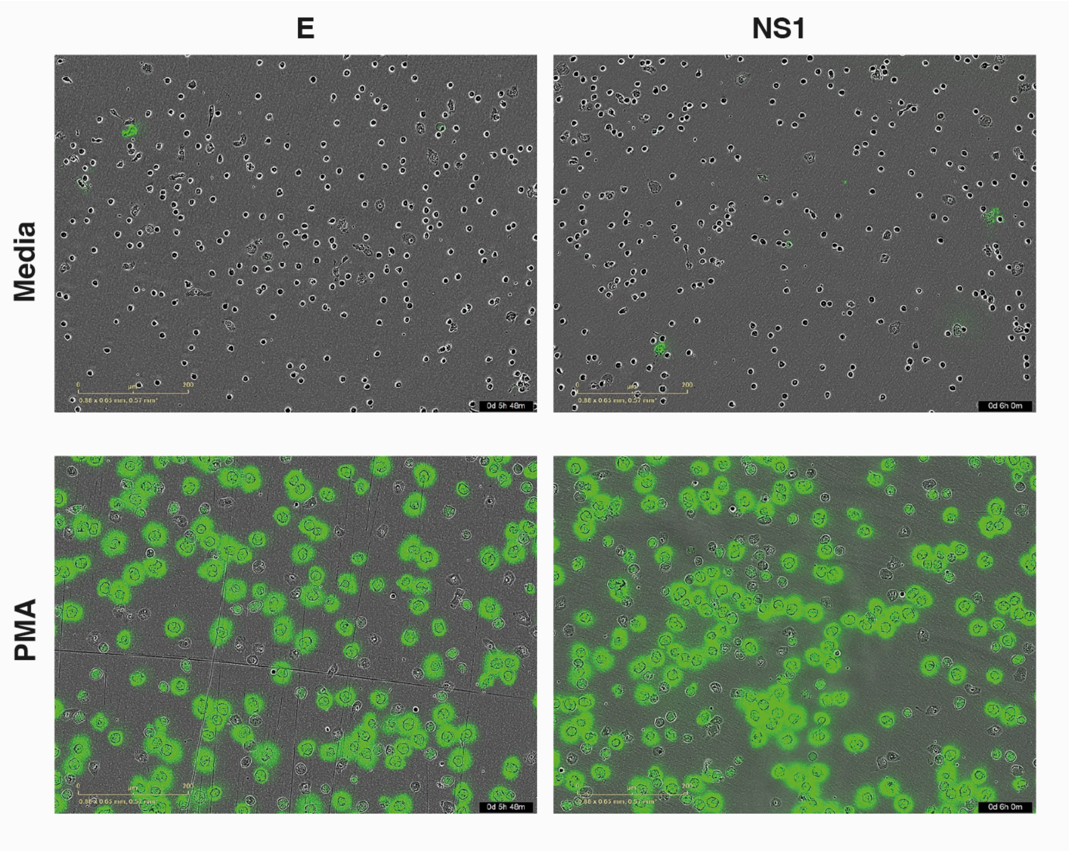


**Extended Data Figure 4. Neutrophil activation and functional *ex vivo* responses to E and NS1 antigens in the presence of pre-infection antibodies.** Representative images of NETosis assays. Neutrophils were incubated with medium alone (negative control) or phorbol 12-myristate 13-acetate (PMA; positive control) in the presence of E or NS1 antigens. Green fluorescence indicates extracellular DNA release characteristic of NET formation. Scale bars: 200 µm.

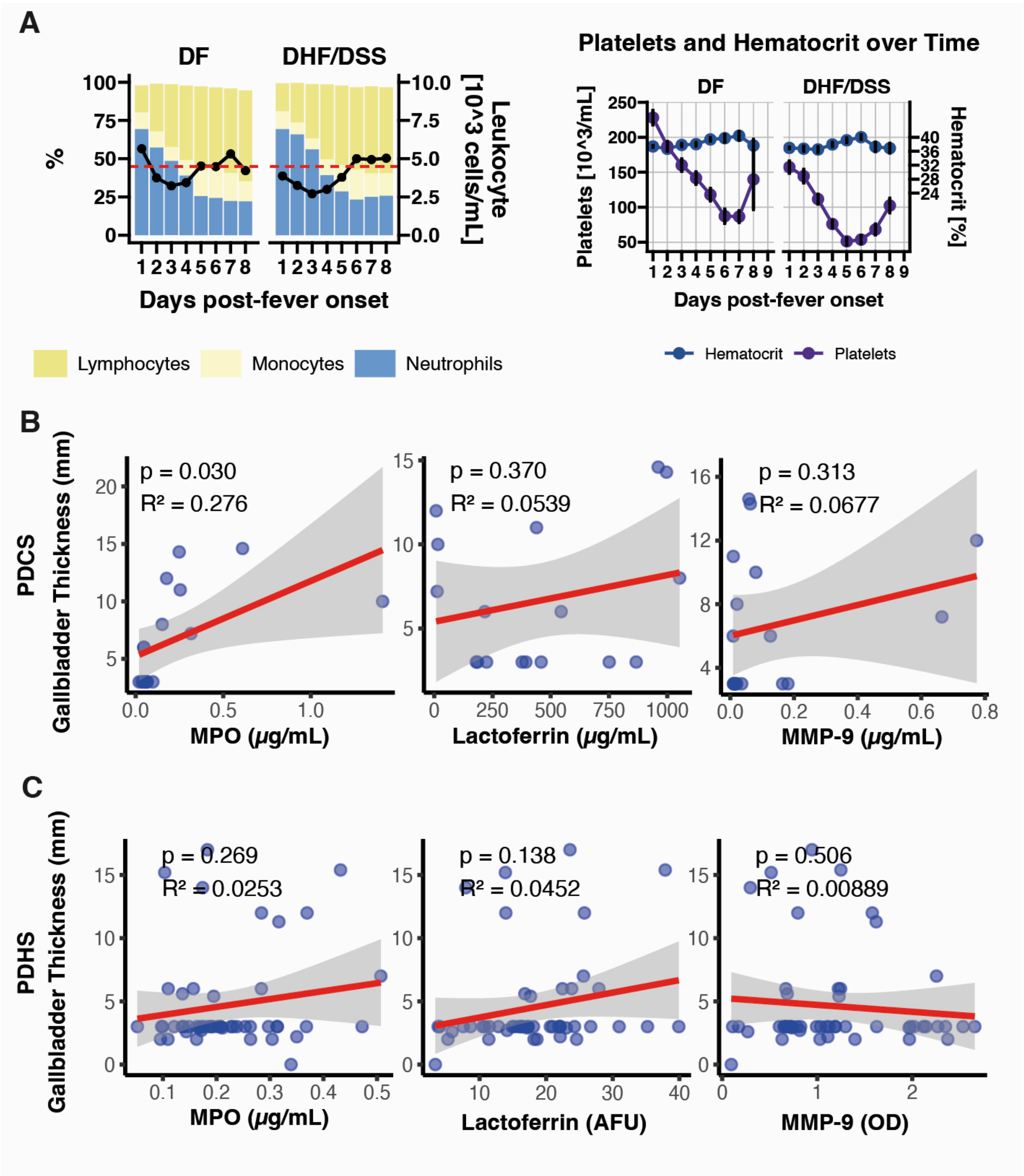


**Extended Data Figure 5. Longitudinal dynamics of leukocyte populations, platelets and hematocrit in the Pediatric Dengue Hospital-based Study (PDHS) and correlations between degranulation markers and gallbladder wall thickness in the Pediatric Dengue Cohort Study (PDCS) and PDHS**. (**A**) Temporal dynamics of leukocyte populations (lymphocytes, monocytes, and neutrophils) in DF and DHF/DSS cases (left panel). The dashed red line marks the cutoff of normal leukocyte counts. Platelet counts and hematocrit trajectories during acute infection are shown (right panel) in the PDHS. (**B**) Pearson's correlation analysis in the PDCS between gallbladder wall thickness and neutrophil degranulation markers, including myeloperoxidase (MPO), lactoferrin, and matrix metalloproteinase-9 (MMP-9). Raw p-values (non adjusted by multiple comparison) are displayed. MPO showed a significant positive correlation with gallbladder wall thickness ($p = 0.030$, $R^2 = 0.275$). (**C**) Correlation analysis in the PDHS between gallbladder wall thickness and neutrophil degranulation markers.

| Outcome | n | % Male | Mean Days | Mean Age | Leukocyte count | | Platelet count | |
|---|---|---|---|---|---|---|---|---|
| | | | | | Mean | SD | Mean | SD |
| **Pediatric Dengue Cohort study** | | | | | | | | |
| DF | 14 | 42.86 | 1.5 | 10.44 | 3.31 | 0.88 | 129.57 | 40.59 |
| DHF/DSS | 10 | 40.00 | 1.9 | 10.67 | 2.94 | 0.83 | 43.20 | 21.34 |
| **Pediatric Dengue Hospital study** | | | | | | | | |
| DF | 36 | 47.22 | 2.61 | 9.41 | 3.02 | 0.96 | 127.14 | 55.73 |
| DHF/DSS | 18 | 44.44 | 2.39 | 10.49 | 2.41 | 0.62 | 49.16 | 28.95 |

**Table S1.** Clinical and laboratory characteristics of pediatric dengue cases in the cohort and hospital-based studies. Summary of demographic, clinical, and laboratory features of children with dengue fever (DF) or dengue hemorrhagic fever/dengue shock syndrome (DHF/DSS) in the Pediatric Dengue Cohort Study and Pediatric Dengue Hospital Study. For each outcome group, the table presents the total number of cases (n), percentage of male participants, mean time to sample collection post-fever onset (days), mean age (in years), mean leukocyte count ($10^3/\mu L$) with standard deviation (SD), and mean platelet count ($10^3/\mu L$) with standard deviation (SD).
